# Personalised risk prediction tools for cryptococcal meningitis mortality to guide treatment stratification; a pooled analysis of two randomised-controlled trials

**DOI:** 10.1101/2024.07.10.24310212

**Authors:** T.H.A. Samuels, S.F. Molloy, D.S. Lawrence, A. Loyse, C. Kanyama, R.S. Heyderman, W.S. Lai, S. Mfinanga, S. Lesikari, D. Chanda, C. Kouanfack, E. Temfack, O. Lortholary, M.C. Hosseinipour, A.K. Chan, D.B. Meya, D.R. Boulware, H.C. Mwandumba, G. Meintjes, C. Muzoora, M. Mosepele, C.E. Ndhlovu, N. Youssouf, T.S. Harrison, J.N. Jarvis, R.K. Gupta

**Author notes:** Correspondence Dr Rishi K Gupta, UCL Respiratory, 1^st^ Floor Rayne Building, 5 University Street, London, WC1E 6JF, United Kingdom. Contributed equally as first authors (T.H.A. Samuels, S.F. Molloy). Contributed equally as senior authors (T.S. Harrison, J.N. Jarvis, R.K. Gupta). ***Table of authors:*** Thomas HA Samuels Dr. MRCP, Sile F Molloy Dr. Ph.D., David S Lawrence Dr. Ph.D., Angela Loyse Dr. M.D.(Res.), Cecilia Kanyama Dr. M.D., Robert S Heyderman Professor FMedSci, Wai Shing Lai Mr. BSc, Sayoki Mfinanga Professor Ph.D., Sokoine Lesikari Dr M.D., Duncan Chanda Dr. Ph.D., Charles Kouanfack Dr. Ph.D., Elvis Temfack Dr. M.D., Olivier Lortholary Professor Ph.D., Mina C Hosseinipour Professor M.D., Adrienne K Chan Dr. M.D., David B Meya Dr. Ph.D., David R Boulware Professor M.D., Henry C Mwandumba Professor Ph.D., Graeme Meintjes Professor Ph.D., Conrad Muzoora Dr. M.D., Mosepele Mosepele Professor. M.D., Chiratidzo E Ndhlovu Professor F.R.C.P., Nabila Youssouf Dr. Ph.D., Thomas S Harrison Professor M.D., Joseph N Jarvis Professor Ph.D., Rishi K Gupta Dr. Ph.D.

## Abstract

**Background:** Cryptococcal meningitis is a leading cause of adult community-acquired meningitis in sub-Saharan Africa with high mortality rates in the first 10 weeks post diagnosis. Practical tools to stratify mortality risk may help to tailor effective treatment strategies.

**Methods:** We pooled individual-level data from two randomised-controlled trials of HIV-associated cryptococcal meningitis across eight sub-Saharan African countries (ACTA, ISRCTN45035509; Ambition-cm, ISRCTN72509687). We used this pooled dataset to develop and validate multivariable logistic regression models for 2-week and 10-week mortality. Candidate predictor variables were specified *a priori*. ‘Basic’ models were developed using only predictors available in resource-limited settings; ‘Research’ models were developed from all available predictors. We used internal-external cross-validation to evaluate performance across countries within the development cohort, before validation of discrimination, calibration and net benefit in held-out data from Malawi (Ambition-cm trial). We also evaluated whether treatment effects in the trials were heterogenous by predicted mortality risk.

**Findings:** We included 1488 participants, of whom 236 (15.9%) and 469 (31.5%) met the 2-week and 10-week mortality outcomes, respectively. In the development cohort (n=1263), five variables were selected into the basic model (haemoglobin, neutrophil count, Eastern Cooperative Oncology Group performance status, Glasgow coma scale and treatment regimen), with two additional variables in the research model (cerebrospinal fluid quantitative culture and opening pressure) for 2-week mortality. During internal-external cross-validation, both models showed consistent discrimination across countries (pooled areas under the receiver operating characteristic curves (AUROCs) 0.75 (95% CI 0.68-0.82) and 0.78 (0.75-0.82) for the ‘Basic’ and ‘Research’ 2-week mortality models, respectively), with some variation in calibration between sites. Performance was similar in held-out validation (n=225), with the models demonstrating higher net benefit to inform decision-making than alternative approaches including a pre-existing comparator model. In exploratory analyses, treatment effects varied by predicted mortality risk, with a trend towards lower absolute and relative mortality for a single high-dose liposomal Amphotericin B-based regimen (in comparison to 1-week Amphotericin B deoxycholate plus flucytosine) among lower risk participants in the Ambition-cm trial.

**Interpretation:** Both models accurately predict mortality, were generalisable across African trial settings, and have potential to be incorporated into future treatment stratification approaches in low and middle-income settings.

**Funding:** MRC, United Kingdom (100504); ANRS, France (ANRS12275); SIDA, Sweden (TRIA2015-1092); Wellcome/MRC/UKAID Joint Global Health Trials (MR/P006922/1); European DCCT Partnership; NIHR, United Kingdom through a Global Health Research Professorship to JNJ (RP-2017-08-ST2-012) and a personal Fellowship to RKG (NIHR302829).

**RESEARCH IN CONTEXT:** *Evidence before this study:* There is an urgent need to improve clinical management for HIV-associated cryptococcal meningitis in resource limited settings across Africa. Cryptococcal meningitis accounts for ∼112,000 AIDS-related deaths per year globally, with over 75% in Africa, despite widespread antiretroviral therapy roll-out. The development of practical tools to identify patients at highest risk of death could help to tailor management strategies and stratify therapy. We searched PubMed for studies published between database inception and Jan 12, 2024, using the terms “cryptococcal meningitis”, “HIV”, “human immunodeficiency virus”, “immunocompromised”, “predict*”, and “model*”, with no language restrictions. Three previous studies, all conducted in China, have developed prognostic models for cryptococcal meningitis mortality. Of these, two used statistical methods while the third used machine learning but focused on persons without HIV only. No studies conducted in Africa, specifically targeting people living with HIV, or using both statistical and machine learning approaches in parallel, were identified. Well-developed and validated tools to predict risk of cryptococcal meningitis mortality and guide treatment stratification are thus lacking for resource limited settings in Africa.

*Added value of this study:* To our knowledge, this is the largest study to date to develop and validate prediction models for HIV-associated cryptococcal meningitis mortality. We combined high-quality data from the two largest randomised-controlled clinical trials conducted to date for cryptococcal meningitis treatment, with a total sample size of 1488 participants of whom 236 (15.9%) and 469 (31.5%) met the 2-week and 10-week mortality outcomes, respectively. We developed two models, ‘basic’ and ‘research’, to enable use in both resource-limited and research settings (where additional prognostic markers such as measurements of cerebrospinal fluid (CSF) opening pressure and CSF fungal burden may also be available). In the 2-week mortality models, five variables were included in the ‘basic’ model, with two additional variables included in the ‘research’ model. Both models predicted risk of mortality with consistent discrimination and calibration across sub-Saharan African settings. Head-to-head statistical (logistic regression) and machine learning (XGBoost) methods revealed no added value of the machine learning approach. In exploratory analyses, treatment effects varied by predicted 2-week mortality risk, thus providing proof-of-concept for future treatment stratification approaches. Specifically, there was a trend towards lower mortality for a single high-dose liposomal Amphotericin B-based regimen (in comparison to 1-week Amphotericin B deoxycholate plus flucytosine) among lower risk participants in the Ambition-cm trial.

*Implications of all the available evidence:* The personalised risk predictor for cryptococcal meningitis (PERISKOPE-CM) models accurately predicted mortality risk among patients with HIV-associated cryptococcal meningitis and demonstrated generalisable performance across trial settings in Africa. Predictions from the models could be utilised to direct treatment stratification approaches in future clinical trials, with patients at lowest predicted risk receiving less intensive and less toxic therapy. The models have been made available for future research use on an open access online interface.

## INTRODUCTION

Cryptococcal meningitis is a major driver of global HIV-related mortality resulting in ∼112,000 deaths per year, with over 75% in Africa, and accounting for 19% of all AIDS-related deaths.^1^ Despite the rapid roll-out of antiretroviral therapy (ART), incidence remains high and ten-week mortality ranges from 24% to over 50%, depending on setting and treatment used.^2–7^ Stratifying patients by disease severity on admission may allow for more targeted therapy; less sick patients could avoid both prolonged hospitalisation and the most intensive and toxic treatments, whereas those who are severely unwell could be identified early for treatment escalation. Validated approaches to prognostication could be used in interventional trials to direct treatment stratification.

Previous studies have identified factors associated with increased risk of HIV-associated cryptococcal meningitis mortality including older age,^8–10^ clinical measurements at presentation (such as low weight/body mass index (BMI),^8,11,12^ abnormal mental status,^8,9,12–15^ and CSF opening pressure^8,9^), and laboratory measurements (including low cerebrospinal fluid (CSF) white cell count (WCC),^12–14,16^ elevated CSF fungal burden,^8,10,12,13,15^ high peripheral white blood cell (WBC) count,^8^ low CD4 cell count,^9^ low haemogloblin,^8^ and high serum C-reactive protein^17^). However, the majority of studies published to date are small, with few conducted in Africa.^8,11^ There are currently no practical tools leveraging the combined discrimination of these factors to identify those at highest risk of death in clinical use. Prognostic models have been developed for a range of acute infectious diseases, notably COVID-19,^18,19^ where extensive external validation has been performed for a model to predict in-hospital mortality. For cryptococcal meningitis, existing prognostic models have been developed in studies from China using conventional statistical methods^9,20^ and for persons without HIV using machine learning approaches.^21^ However, there are currently no validated models to guide clinical decision-making in people living with advanced HIV in Africa, where the disease burden is highest.

In this study, we pooled individual-level data from the two largest randomised-controlled trials (RCTs) to date in HIV-associated cryptococcal meningitis. We aimed to develop and validate prediction models to predict risk of all-cause 2-week mortality. We used both statistical and machine learning approaches, in a head-to-head analysis, and also evaluated performance over a longer (10-week) time horizon. Finally, we assessed whether treatment efficacy in the trials was heterogenous by predicted risk.

## METHODS

### Data sources

We analysed pooled data from the ACTA (ISRCTN45035509) and Ambition-cm (ISRCTN72509687) phase III, RCTs for HIV-associated cryptococcal meningitis, which have been reported previously.^4,5^

In brief, adults (≥18 years) with a first episode of HIV-associated cryptococcal meningitis were recruited to both trials. In ACTA, participants were randomized in a 1:1:1 ratio to received either: fluconazole plus flucytosine for 14 days (oral combination regimen); amphotericin B deoxycholate plus either flucytosine or fluconazole for 7 days followed by 7 days fluconazole; or amphotericin B deoxycholate plus either flucytosine or fluconazole for 14 days. Participants were recruited from 9 sites across 4 countries in Africa (Cameroon, Malawi, Tanzania and Zambia). In Ambition-cm, participants were randomized 1:1 to receive either: single high-dose (10 mg/kg) liposomal amphotericin B plus 14 days of flucytosine plus fluconazole (Ambition regimen); or amphotericin B deoxycholate (1 mg/kg/day) plus flucytosine for 7 days, followed by fluconazole for 7 days (1-wk AmBd+5FC regimen). Participants were recruited from 6 sites in 5 countries (Botswana, Malawi, South Africa, Uganda and Zimbabwe).

We elected *a priori* to include all data in model development apart from the Malawi site in the Ambition-cm trial, which was held-out for validation since Malawi was the only country where participants were recruited to both trials.

Both studies were approved by the London School of Hygiene and Tropical Medicine Research Ethics Committee and local ethics and regulatory authorities in each country, where appropriate. The full analysis pipeline is summarised in Supplementary Figure 1. All analyses were conducted and reported in accordance with TRIPOD standards^7^ and were performed in R (version 4.3.2).

### Model development

We considered predefined candidate predictors for inclusion in the models based on clinical knowledge, previous studies and availability of variables collected at baseline in both trials (Supplementary Table 1). Sample size calculation details are provided in the Supplementary Appendix. Variables were only considered if available from at least 60% of participants, as previously.^23^ Missing data were handled using multiple imputation in the primary analyses, using the aRegImpute function in R.^23,24^ All predictors, including transformations, considered for the final model were included in the imputation model to ensure compatibility. All primary analyses were performed across 10 multiply imputed datasets; parameters were pooled using Rubin’s rules.^25^

We developed models for the primary outcome of 2-week mortality, since we hypothesised that early mortality is more likely to be directly associated with cryptococcal meningitis severity. We developed a ‘basic’ model for use in a resource limited setting, using only candidate predictors routinely obtained programmatically at the point of hospital admission; and a ‘research’ model, where all pre-defined candidate predictors were considered for inclusion. Multivariable logistic regression models were initially developed in the development dataset. Variable selection was conducted for the ‘basic’ and ‘research’ models separately, using backward selection, based on Akaike Information Criteria (AIC). Continuous variables were modelled using restricted cubic splines (3 knots) to assess non-linear associations. Predictors retained in >50% of multiply imputed sets were retained in the final models.

Models including the selected variables were then evaluated using the internal-external cross-validation method, where participants from one country were iteratively left out of the model development set and used for validation.^23,26,27^ This method evaluates potential generalisability of the modelling approach between settings by examining between-setting heterogeneity in performance. To do this, the models were re-trained in the remaining countries in the development dataset and validated in the omitted country by quantifying the discrimination and calibration. Discrimination assesses how well the models differentiate between those who do and do not meet the outcome (measured as the C-statistic), while calibration evaluates how well predicted risk matches observed risk (measured as the calibration slope and calibration-in-the-large, and by visualisation of calibration plots^28^). Random-effects meta-analysis was then used to calculate pooled measures of discrimination and calibration across countries in the development dataset.^26^ We performed re-calibration by country by re-estimating model intercepts. The final models were trained using the full development dataset prior to further validation in the held-out dataset (Malawi site in the Ambition-cm trial).

### Machine learning

Next, we sought to test whether a statistical model using logistic regression could be improved upon using a machine learning method. We re-trained the research model with the selected variables using XGBoost, as a “best-in-class” machine learning approach for predicting a binary outcome,^29^ as previously.^19^ Further details of the XGBoost approach are provided in the Supplementary Appendix.

### Model validation

Held-out validation of the statistical and machine learning models was then performed in the Ambition-cm Malawi dataset by quantifying the C-statistic, calibration slope and calibration-in-the-large, and visualisation of calibration plots. We benchmarked performance to single univariable predictors, and other HIV-associated cryptococcal meningitis prognostic models for which constituent variables were available in >60% of the dataset and model reconstruction was possible from reported manuscripts.^9,30^ We also performed decision curve analysis^30^ in the validation dataset to quantify the overall net benefit of implementing the models to inform clinical decisions, compared to: (a) a ‘treat all’ approach; (b) a ‘treat none’ approach; and (c) other candidate models.

### 10-week analysis

Performance of the final logistic regression models to predict mortality over a longer time horizon of 10 weeks was assessed by evaluating discrimination for this outcome in the held-out validation in the Malawi Ambition-cm dataset. In addition, we developed separate models specifically to predict the 10-week outcome, using the same methodology as for the primary models, to assess whether this further improved performance for the longer time horizon.

### Treatment effect heterogeneity analyses

In an exploratory analysis, we also tested the hypothesis that the Ambition regimen and oral combination (fluconazole plus flucytosine) regimen were more effective than 1-wk AmBd+5FC (as the control arm in Ambition-cm and the best performing arm in ACTA), among lower risk participants, in line with the Predictive Approaches to Treatment effect Heterogeneity (PATH) Statement.^31^ To do this, we generated predictions for 2-week mortality using the final ‘basic’ and ‘research’ models in the pooled dataset.

Participants were divided into low, medium and high-risk strata (based on tertiles of predicted risk). We then examined treatment efficacy of the Ambition and oral combination regimens (compared to 1-wk AmBd+5FC) over 10 weeks separately within each trial, with interaction terms by risk stratum. Absolute risk differences between arms were quantified by risk stratum using an identity link function and relative risk as hazard ratios using Cox regression.

In parallel, we evaluated whether treatment efficacy was modified by predicted risk as a continuous variable. We modelled time to death using Cox regression, with an interaction term between treatment arm and predicted 2-week mortality risk (including restricted cubic spline transformations to account for non-linear associations). We then visualised treatment effects (as hazard ratios) against predicted risk.

## RESULTS

### Baseline characteristics

A total of 674 eligible participants from the ACTA trial (excluding 4 participants who were lost to follow-up) and 814 from the Ambition-cm trial were included in the pooled analysis (n=1,488). 1,263 participants were included in model development, with 225 (Malawi Ambition-cm participants) held out for validation (Supplementary Figure 2). Baseline characteristics, including candidate predictors and missingness, are presented in Table 1, stratified by 2-week mortality. A total of 222/1,263 (17.5%) development set participants and 21/225 (9.3%) validation set participants met the primary model outcome of 2-week mortality.

**Table 1:**
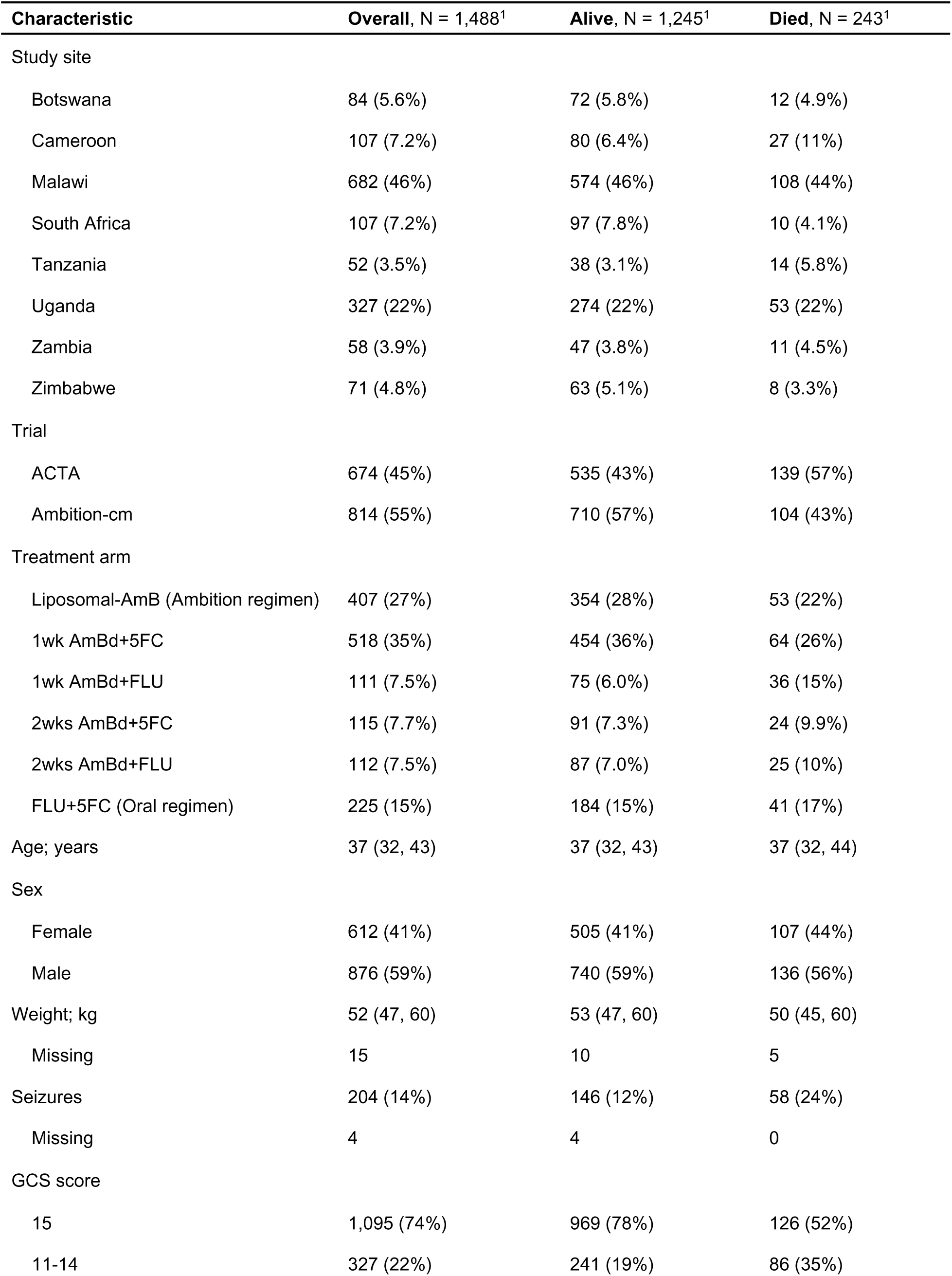
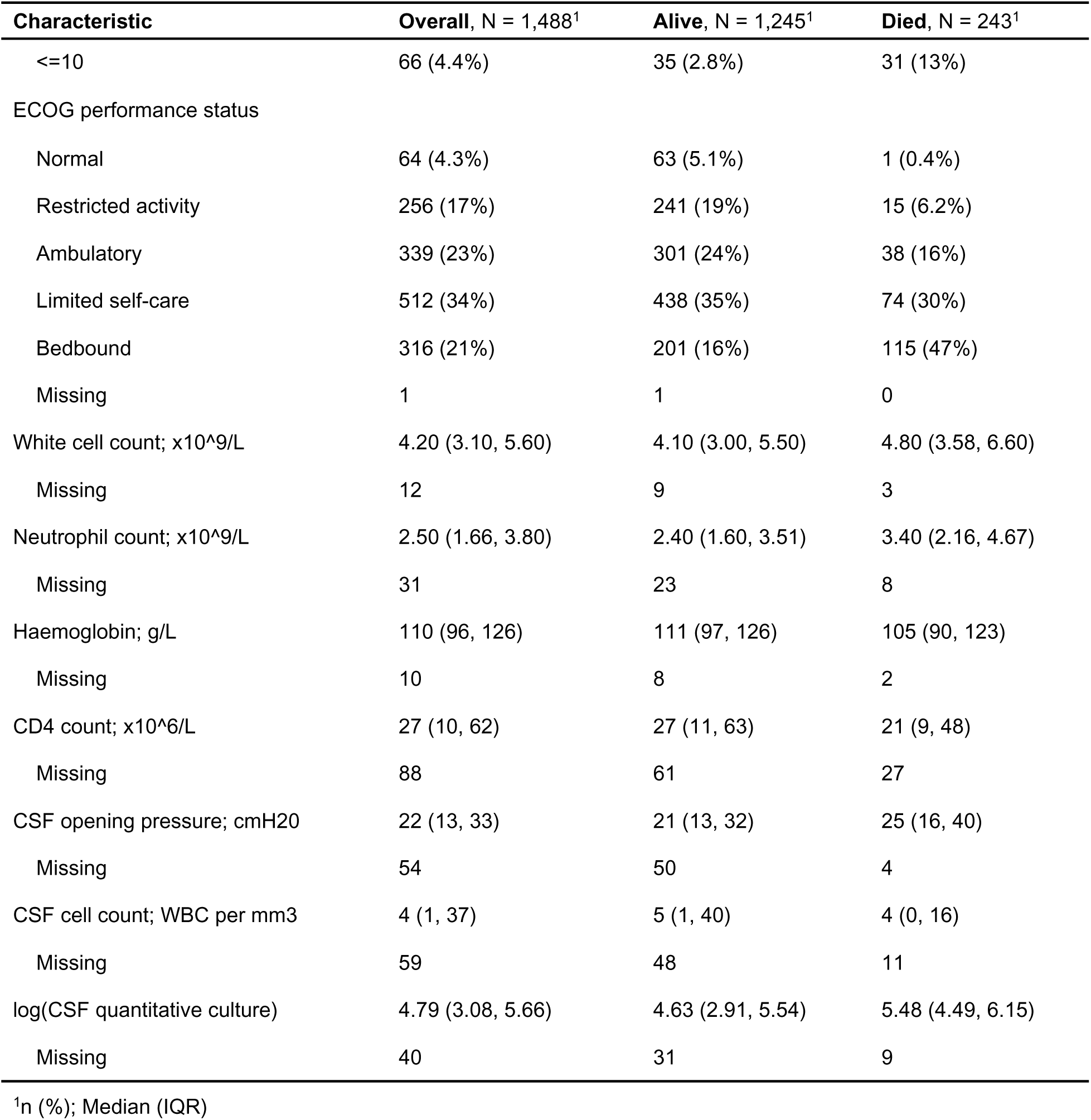
Baseline characteristics of study cohort. Characteristics showed both in total study cohort and stratified by two-week mortality outcome.

| Characteristic | Overall, N = 1,488<sup>1</sup> | Alive, N = 1,245<sup>1</sup> | Died, N = 243<sup>1</sup> |
| --- | --- | --- | --- |
| Study site |  |  |  |
| Botswana | 84 (5.6%) | 72 (5.8%) | 12 (4.9%) |
| Cameroon | 107 (7.2%) | 80 (6.4%) | 27 (11%) |
| Malawi | 682 (46%) | 574 (46%) | 108 (44%) |
| South Africa | 107 (7.2%) | 97 (7.8%) | 10 (4.1%) |
| Tanzania | 52 (3.5%) | 38 (3.1%) | 14 (5.8%) |
| Uganda | 327 (22%) | 274 (22%) | 53 (22%) |
| Zambia | 58 (3.9%) | 47 (3.8%) | 11 (4.5%) |
| Zimbabwe | 71 (4.8%) | 63 (5.1%) | 8 (3.3%) |
| Trial |  |  |  |
| ACTA | 674 (45%) | 535 (43%) | 139 (57%) |
| Ambition-cm | 814 (55%) | 710 (57%) | 104 (43%) |
| Treatment arm |  |  |  |
| Liposomal-AmB (Ambition regimen) | 407 (27%) | 354 (28%) | 53 (22%) |
| 1wk AmBd+5FC | 518 (35%) | 454 (36%) | 64 (26%) |
| 1wk AmBd+FLU | 111 (7.5%) | 75 (6.0%) | 36 (15%) |
| 2wks AmBd+5FC | 115 (7.7%) | 91 (7.3%) | 24 (9.9%) |
| 2wks AmBd+FLU | 112 (7.5%) | 87 (7.0%) | 25 (10%) |
| FLU+5FC (Oral regimen) | 225 (15%) | 184 (15%) | 41 (17%) |
| Age; years | 37 (32, 43) | 37 (32, 43) | 37 (32, 44) |
| Sex |  |  |  |
| Female | 612 (41%) | 505 (41%) | 107 (44%) |
| Male | 876 (59%) | 740 (59%) | 136 (56%) |
| Weight; kg | 52 (47, 60) | 53 (47, 60) | 50 (45, 60) |
| Missing | 15 | 10 | 5 |
| Seizures | 204 (14%) | 146 (12%) | 58 (24%) |
| Missing | 4 | 4 | 0 |
| GCS score |  |  |  |
| 15 | 1,095 (74%) | 969 (78%) | 126 (52%) |
| 11-14 | 327 (22%) | 241 (19%) | 86 (35%) |
| <=10 | 66 (4.4%) | 35 (2.8%) | 31 (13%) |
| ECOG performance status |  |  |  |
| Normal | 64 (4.3%) | 63 (5.1%) | 1 (0.4%) |
| Restricted activity | 256 (17%) | 241 (19%) | 15 (6.2%) |
| Ambulatory | 339 (23%) | 301 (24%) | 38 (16%) |
| Limited self-care | 512 (34%) | 438 (35%) | 74 (30%) |
| Bedbound | 316 (21%) | 201 (16%) | 115 (47%) |
| Missing | 1 | 1 | 0 |
| White cell count; x10<sup>9</sup>/L | 4.20 (3.10, 5.60) | 4.10 (3.00, 5.50) | 4.80 (3.58, 6.60) |
| Missing | 12 | 9 | 3 |
| Neutrophil count; x10<sup>9</sup>/L | 2.50 (1.66, 3.80) | 2.40 (1.60, 3.51) | 3.40 (2.16, 4.67) |
| Missing | 31 | 23 | 8 |
| Haemoglobin; g/L | 110 (96, 126) | 111 (97, 126) | 105 (90, 123) |
| Missing | 10 | 8 | 2 |
| CD4 count; x10<sup>6</sup>/L | 27 (10, 62) | 27 (11, 63) | 21 (9, 48) |
| Missing | 88 | 61 | 27 |
| CSF opening pressure; cmH2O | 22 (13, 33) | 21 (13, 32) | 25 (16, 40) |
| Missing | 54 | 50 | 4 |
| CSF cell count; WBC per mm3 | 4 (1, 37) | 5 (1, 40) | 4 (0, 16) |
| Missing | 59 | 48 | 11 |
| log(CSF quantitative culture) | 4.79 (3.08, 5.66) | 4.63 (2.91, 5.54) | 5.48 (4.49, 6.15) |
| Missing | 40 | 31 | 9 |

<sup>1</sup>n (%); Median (IQR)

### Variable selection

In variable selection, five predictors were retained in the ‘basic’ and seven in the ‘research’ model. Predictors in both models included: Glasgow coma scale (GCS) score, Eastern Cooperative Oncology Group (ECOG) performance status, haemoglobin, blood neutrophil count and treatment regimen. Additional predictors in the research model were CSF opening pressure (CSF OP) and log10CSF quantitative cryptococcal culture (CSF QCC). Predictor-outcome associations were similar in both models (Figure 1; Supplementary Figure 3). Full model coefficients for both models are presented in Supplementary Tables 4 and 5.

**Figure 1:**
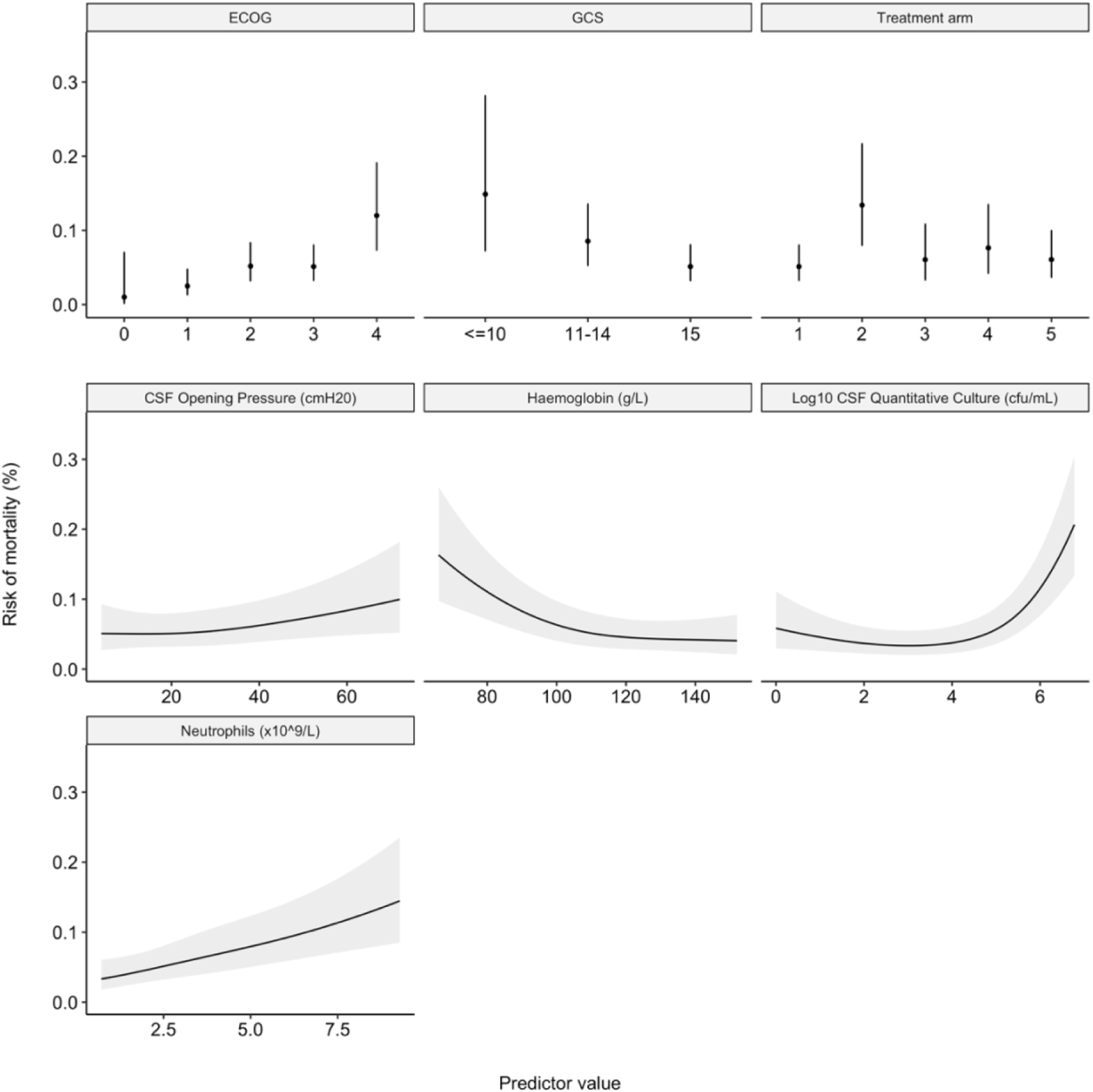
Multivariable associations between selected predictors and outcome in primary research model. Continuous variables were modeled using restricted cubic splines. The final model parameters are pooled across multiply imputed datasets (total sample size for model development = 1,263 participants). For continuous variables, black lines represent point estimates and grey shaded regions represent 95% confidence intervals. For categorical variables, black dots represent point estimates and black lines represent 95% confidence intervals. Treatment arm 1 through 5 represent 1) the liposomal-Amphotericin-B Ambition regimen and the 1-week Amphotericin-B + Flucytosine (1wk AmBd + 5FC) arms from both ACTA and Ambition-cm trials, 2) 1 week Amphotericin-B + Fluconazole, 3) 2 weeks Amphotericin-B + Flucytosine, 4) 2 weeks Amphotericin-B + Fluconazole and 5) Flucytosine + Fluconazole oral combination regimen, respectively. Treatment arm (1) was grouped in this way due to the finding of non-inferiority between the Ambition regimen and 1wk AmBd + 5FC arms in the Ambition-cm trial. Associations for the primary basic model are shown in the appendix (Supp Fig 1). ECOG = Eastern Cooperative Oncology Group performance status; GCS = Glasgow Coma Scale; CSF = cerebrospinal fluid.

### Internal-external cross-validation

Forest plots showing discrimination and calibration metrics of both models from each country in internal-external cross validation in the development dataset are shown in Figure 2. Discrimination was relatively consistent between study sites for both models (pooled C-statistic estimates 0.75 (95%CI 0.68-0.82) for the ‘basic’ and 0.78 (95%CI 0.75-0.82) for the ‘research’ models). Calibration was more heterogeneous, with CITL varying by study site in both models, likely reflecting variation in baseline risk between sites (point estimates -0.45 to 0.57 for the ‘basic’ model, -0.34 to 0.42 for the ‘research’ model). Pooled calibration plots by study site show evidence of systematic overestimation of risk in Malawi and underestimation in Uganda. Recalibration to each study site by re-estimation of the model intercept led to improvement in model calibration, as expected (Supplementary Figure 4).

**Figure 2.**
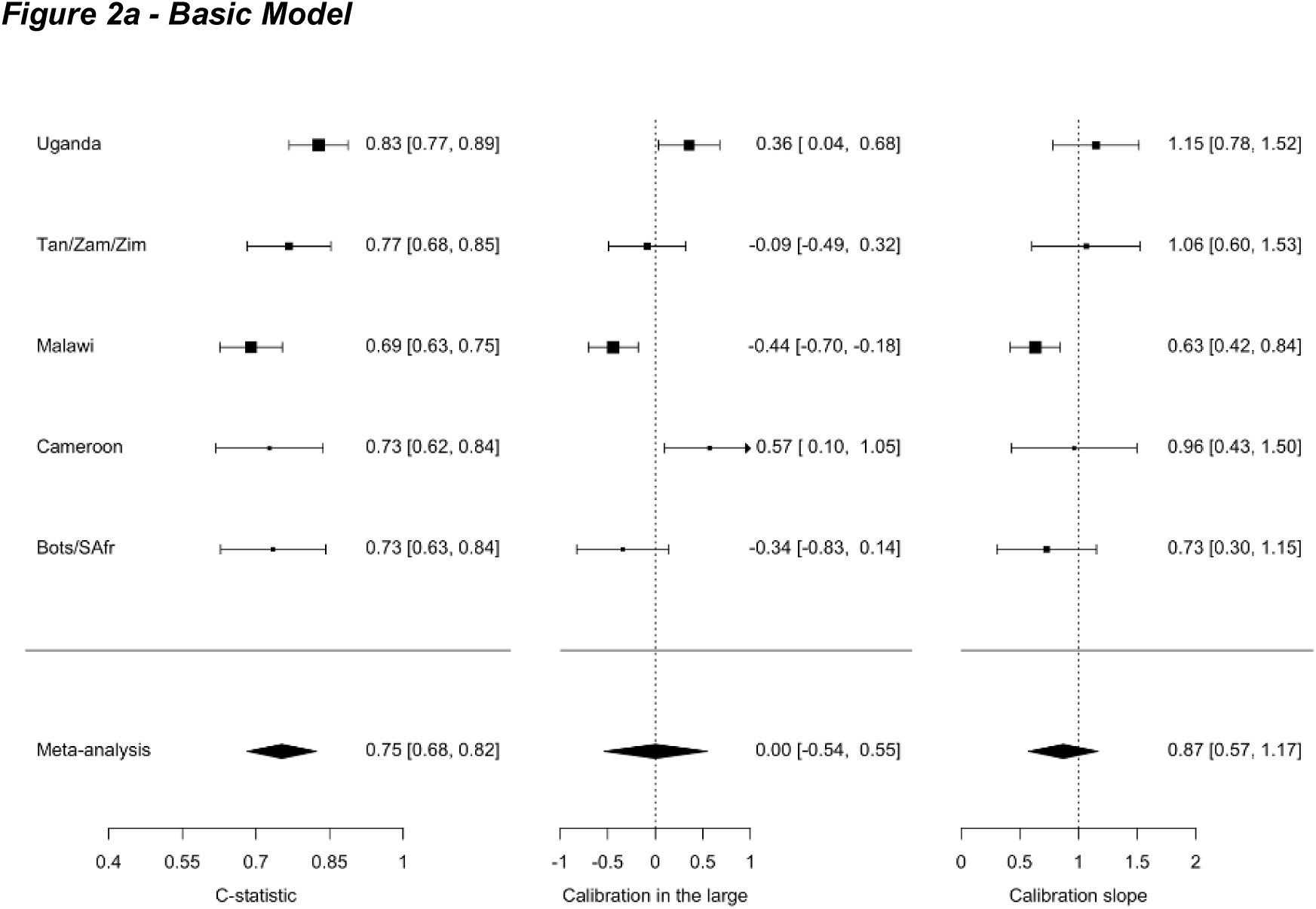
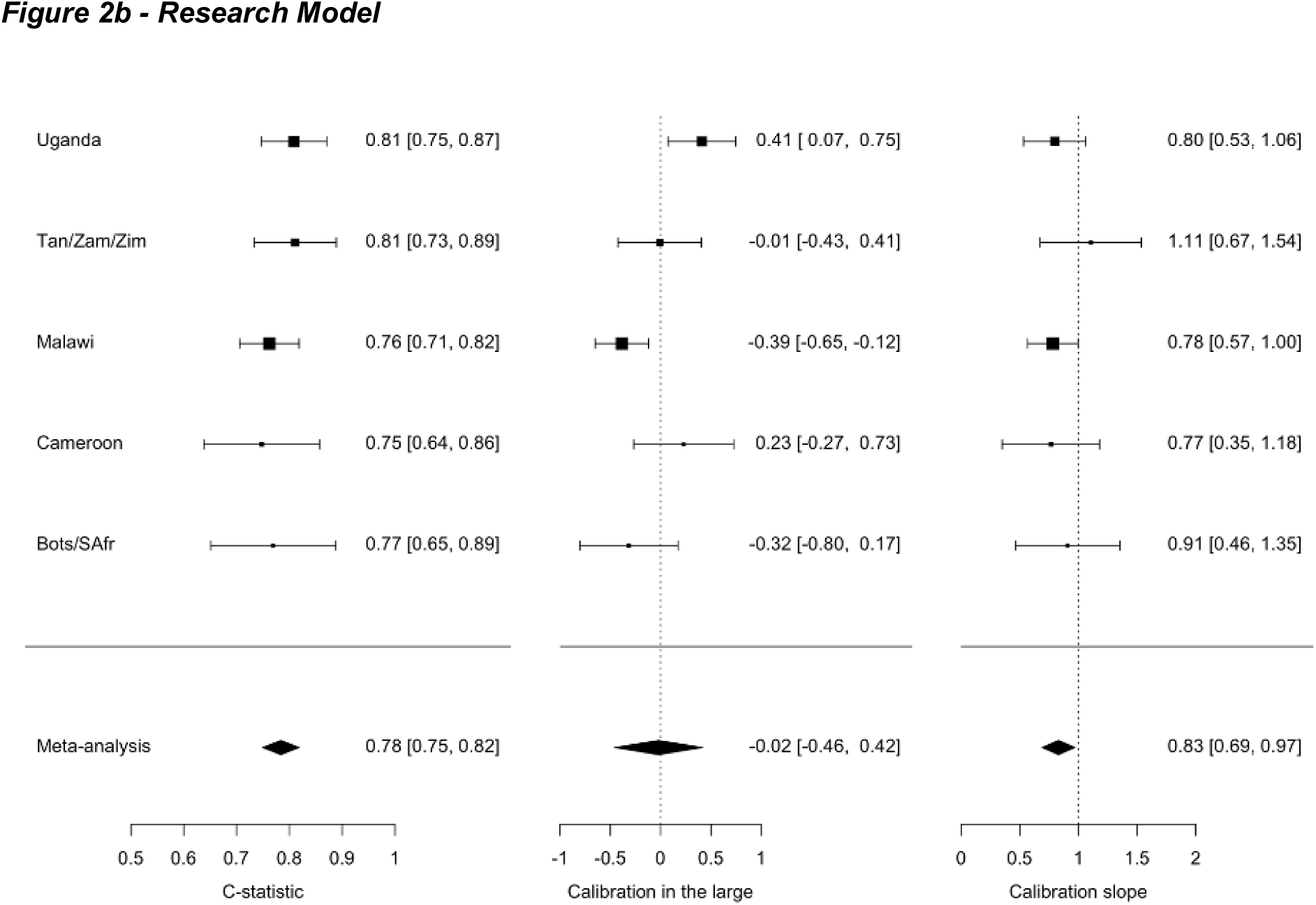
a and 2b: Internal-external cross validation of basic and research model by country. Pooled estimates are calculated through random-effects meta-analysis (total sample size = 1,263 participants). Countries with n < 100 participants or x < 20 deaths were amalgamated and grouped by similarity of healthcare environment. Dashed lines indicate lines of perfect calibration in the large (0) and slope (1), respectively. Black squares indicate point estimates; bars indicate 95% confidence intervals; diamonds indicate pooled random-effects meta-analysis estimates. Bots = Botswana; SAfr = South Africa; Tan = Tanzania; Zam = Zambia; Zim = Zimbabwe.

### Held-out validation

In held-out validation in Malawi participants in the Ambition-cm trial, discrimination of both models was slightly higher compared to the estimates in internal-external cross-validation (Table 2) with C-statistic 0.78 [95%CI 0.70-0.87] and 0.85 [95%CI 0.79-0.92] in the ‘basic’ and ‘research’ model, respectively. Calibration assessment suggested overestimation of risk, particularly in the higher risk range, where data were sparser (Figure 3a-b). ECOG performance status was the strongest univariable predictor for 2-week mortality (C-statistic 0.78 [95%CI 0.71 to 0.85]) but had lower discrimination than the full multivariable ‘research’ model (Supplementary Table 6). The discriminative ability of single predictors varied by study site (Supplementary Table 7). Of the identified pre-existing models, only the model created by Zhao et al. (2021)^9^ could be reconstructed from the available data. Discrimination in the validation dataset, calculated using their reported point score, fell short of either of the primary models (C-statistic 0.69 [95%CI 0.6 to 0.78]). To report calibration, we recalibrated the model intercept to our validation dataset, since no intercept was reported. Despite this, calibration assessment suggested that predictions were too extreme, with a slope 0.35 (95%CI 0.14 - 0.56; Table 2; Supplementary Figure 5).

**Figure 3:**
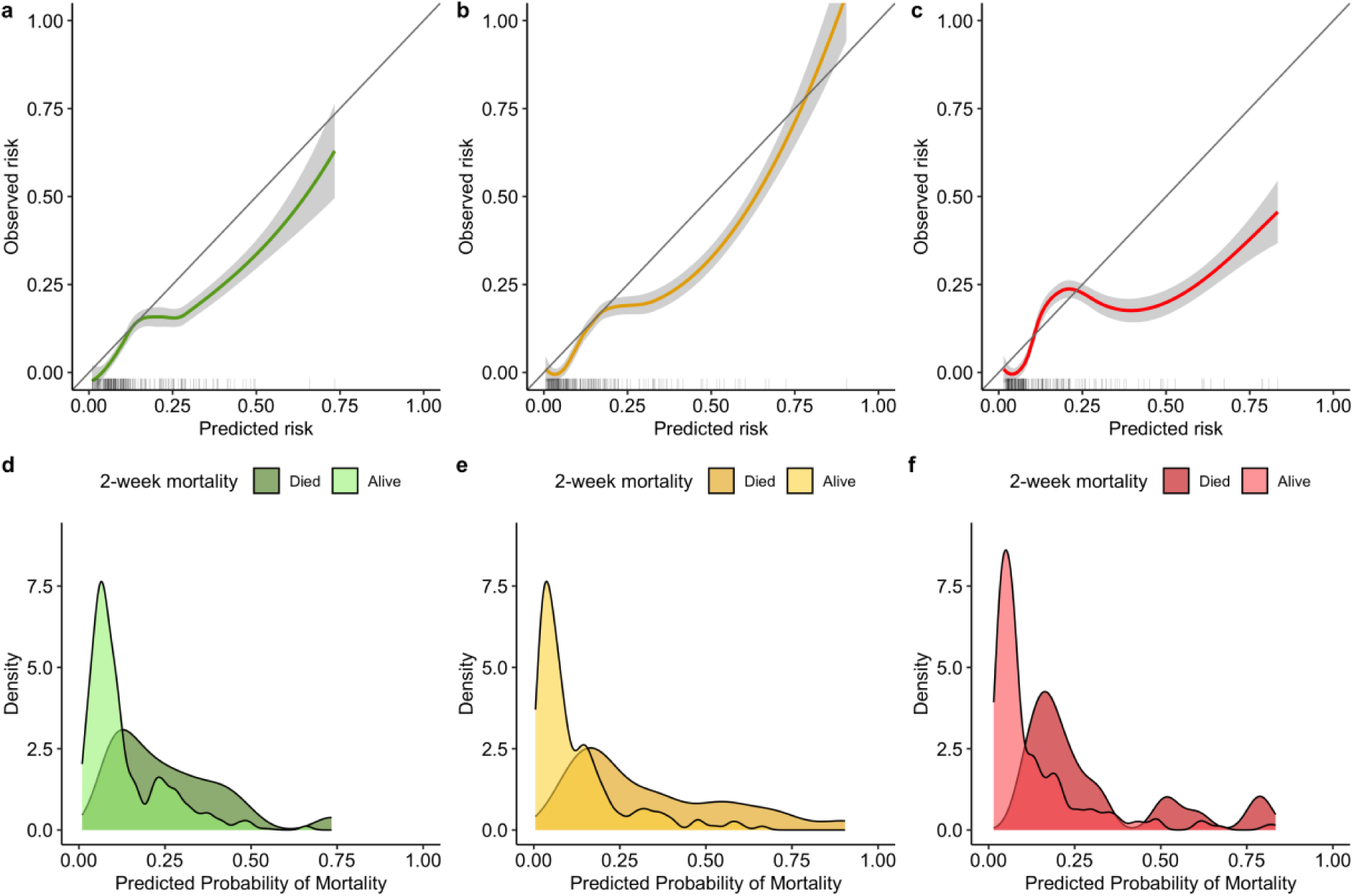
Model calibration and prediction density in held-out validation data. Panels (a) to (c) show the calibration of the basic model, research model and XGBoost machine learning model respectively. Calibration is shown using a loess smoother. 95% confidence intervals are shown shaded in grey. Rug plots, shown on the x-axis, plot the distribution of predicted risk. Panels (d) to (f) show density plots for the 2-week mortality predictions made by the basic model, research model and XGBoost machine learning model respectively, stratified by 2-week mortality outcome.

**Table 2:** Model performance in held-out validation data. Table reports performance in validation data for each of the primary models, secondary models, XGBoost machine learning model and Zhao et al’s model. Brackets in the table headers refer to the outcome for which the model was trained. Validation outcome refers to the outcome in which model performance was assessed. Slope and Calibration-in-the-large data are not reported for the primary models’ 10-week validation outcome, since the primary model was developed for the 2-week mortality outcome. Calibration-in-the-large data is not reported for Zhao et al’s model as in order to reconstruct the model and assess calibration, the model intercept was recalibrated to our data.

|  | Validation Outcome | AUROC<sup>1,2</sup> | Slope<sup>1</sup> | CITL<sup>1,2</sup> |
| --- | --- | --- | --- | --- |
| **Primary models (2-week mortality)** |  |  |  |  |
| Basic | 2 weeks | 0.78 (0.7 - 0.87) | 1.04 (0.54 - 1.55) | -0.55 (-1.02 - -0.07) |
| Basic | 10 weeks | 0.74 (0.66 - 0.81) | - | - |
| Research | 2 weeks | 0.85 (0.79 - 0.92) | 1.14 (0.69 - 1.6) | -0.57 (-1.06 - -0.07) |
| Research | 10 weeks | 0.77 (0.7 - 0.85) | - | - |
| **Secondary models (10-week mortality)** |  |  |  |  |
| Basic | 10 weeks | 0.77 (0.71 - 0.84) | 1.31 (0.85 - 1.77) | -0.16 (-0.48 - 0.16) |
| Research | 10 weeks | 0.78 (0.71 - 0.85) | 1.1 (0.72 - 1.49) | -0.07 (-0.4 - 0.25) |
| **Machine learning model (2-week mortality)** |  |  |  |  |
| XGBoost | 2 weeks | 0.83 (0.76 - 0.89) | 0.87 (0.49 - 1.26) | -0.63 (-1.12 - -0.13) |
| **Zhao et al. model (28-day mortality)** |  |  |  |  |
| Zhao | 28 days | 0.69 (0.6 - 0.78) | 0.35 (0.14 - 0.56) | - |

<sup>1</sup>Brackets indicate 95% confidence intervals
<sup>2</sup>AUROC = Area under the Receiver Operator Characteristic curve; CITL = Calibration-in-the-large

### Machine learning

We also developed an XGBoost model, to assess whether a machine learning approach could further improve performance. Predictor-outcome associations using XGBoost were similar to the logistic regression approach, with no clear evidence of two-way interactions (Supplementary Figure 7). In the validation dataset, XGBoost discrimination and calibration metrics were also similar to the logistic regression model (Table 2; Figure 3). The XGBoost calibration plot showed miscalibration with predictions being too extreme, reflecting overfitting, and leading to a slope <1 (Figure 3c). Overall, performance of the machine learning model demonstrated no improvement in performance over the logistic regression approach.

### Decision curve analysis

Decision curve analysis measures the trade-off between correctly identifying participants who met the mortality endpoint and incorrectly identifying those who did not and quantifies this as “net benefit”^30^. Decision-curve analysis in the validation dataset showed higher net-benefit for the ‘basic’ and ‘research’ models compared with the model by Zhao et al. (2021),^9^ ‘treat-all’ or ‘treat-none’ approaches across a broad range of threshold probabilities, where the weighting of the false positives is varied. The ‘research’ model demonstrated superior net benefit overall to guide management (Supplementary Figure 6).

### 10-week mortality

A total of 411/1263 (32.5%) and 58/225 (26.2%) participants met the 10-week mortality outcome in the development and validation datasets, respectively. We retrained the logistic regression models, including variable selection, to assess whether this would improve prediction over the longer time period. Seven predictors were retained in the retrained ‘basic’, and eight in the ‘research’ 10-week models. In the ‘basic’ model, age, weight, and the presence of seizures on admission were substituted for GCS, and in the ‘research’ model age and CSF cell count were substituted for CSF OP (Supplementary Tables 8-10; Supplementary Figure 9). Performance of the 10-week models in the validation dataset is shown in Table 2, along with the C-statistic for the primary 2-week models in predicting 10-week mortality. Overall, for both ‘basic’ and ‘research’ models, discrimination for the 10-week outcome was similar for the re-trained 10-week models and the primary 2-week models. For both the ‘basic’ and ‘research’ 10-week models, the CITL was closer to 0 for the 10-week outcome, when compared to the primary 2-week models for the 2-week outcome (Supplementary Figure 10), suggesting better calibration for the 10-week models.

### Treatment effect modification analysis

We hypothesised that treatment effects within each trial may vary according to predicted 2-week mortality risk. Predicted risk for both models was positively skewed, with modal risk of 7.6% and 4.2% for the ‘basic’ and ‘research’ models, respectively, markedly lower than the overall cohort mortality of 16.3% (Supplementary Figure 11). Absolute number of deaths, mortality risk difference and hazard ratios comparing the Ambition regimen to 1-wk AmBd+5FC in the Ambition-cm trial and oral combination treatment to 1-wk AmBd+5FC in the ACTA trial, stratified by risk stratum, are shown in Supplementary Table 11. In the Ambition-cm trial, there was a trend towards absolute and relative risk reductions for the Ambition regimen for the lower risk strata for both models, with no difference in mortality in the higher risk stratum: Hazard ratio 0.65 (95%CI 0.38-1.13) for ‘basic’ model in low-risk stratum, compared to 1.05 (95%CI 0.71-1.54) in high-risk stratum; 0.61 (95%CI 0.33-1.11) for ‘research’ model in low-risk stratum, compared to 1 (95%CI 0.68-1.45) in high-risk stratum).

When comparing oral combination treatment to 1-wk AmBd+5FC in the ACTA trial, risk differences and hazard ratios appeared closer to 1 in the low- and medium-risk strata, compared to the high-risk strata, for both models, though the confidence intervals were wide due to small sample sizes in each stratum. These findings were similar when handling predicted risk as a continuous variable, shown in Figure 4.

**Figure 4:**
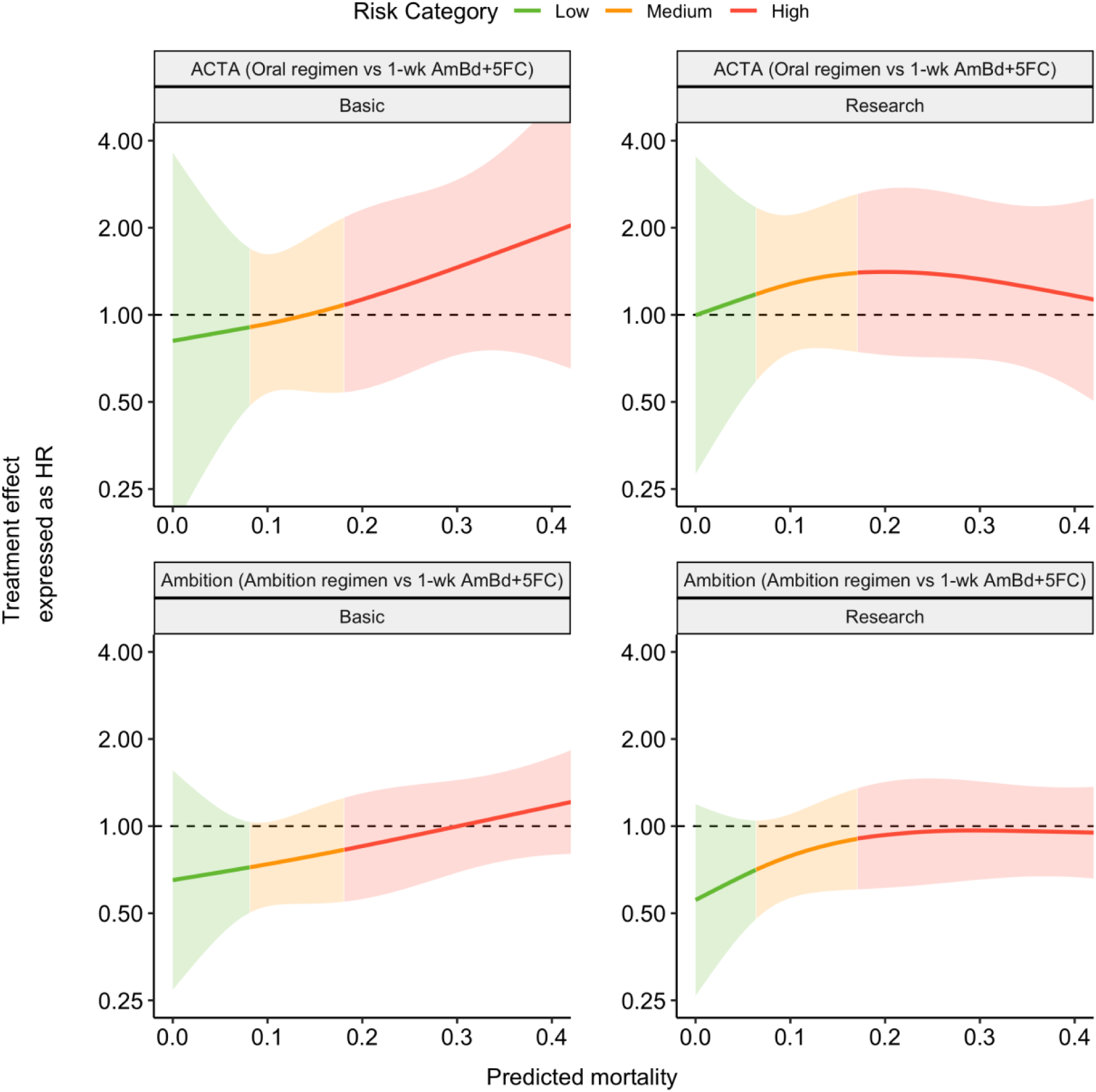
Treatment Effect of Oral regimen (flucytosine (5FC) plus fluconazole (FLU)) and Ambition regimen vs 1-week Amphotericin B deoxycholate (AmBd) plus 5FC over a range of treatment model-predicted mortality. Treatment effect, expressed as a hazard ratio for the Oral or Ambition regimens relative to 1-week of Amphotericin B plus Flucytosine (1wk AmBd + 5FC), is plotted logarithmically to base 2 on the y axis. Predicted mortality, defined by either the Basic or Research model as per the panel labels, is plotted on the x axis. Green, Orange and Red colouring represent the Low, Medium and High Risk terciles defined by the research model. Shaded regions represent 95% confidence intervals. The x-axis is foreshortened to predicted mortality of 0.4, which encompasses >95% of the underlying data.

## DISCUSSION

Using a large, high-quality dataset of the 2 largest randomised controlled trials conducted to date for the treatment of cryptococcal meningitis, conducted across 8 countries in Africa, we developed tools to predict 2-week and 10-week mortality. Our ‘basic’ model includes five predictor variables, which are routinely measured programmatically: GCS score, ECOG performance status, haemoglobin, neutrophil count and treatment regimen. Our ‘research’ model additionally includes CSF OP and CSF fungal burden, for settings where these variables are available. Both models demonstrated consistent performance across countries, supporting their generalisability. Predicted risk was markedly skewed with modal mortality risk far below overall risk, suggesting that the majority of patients have individual-level risk far below the mean and thus supporting an individualised approach to treatment. Moreover, our results provide proof of concept data for treatment stratification. While the Ambition regimen was non-inferior to 1-wk AmBd+5FC overall in the Ambition-cm trial, we observed a trend towards improved mortality with the Ambition regimen among lower risk participants, when using both categorical and continuous risk measurements. Treatment regimens in cryptococcal meningitis are intensive and use of Amphotericin B deoxycholate is associated with significant toxicity risk.^32^ One potential hypothesis is that treatment toxicity may be a relatively more important factor determining outcome among lower-risk participants, which may explain the apparent improved outcomes with the less toxic Ambition regimen^5^ among these participants. When evaluating oral combination therapy in the ACTA trial, the oral regimen was close to the efficacy of 1-wk AmBd+5FC in the lowest risk participants. Our results suggest our models could be used to direct stratified treatment approaches in future trials, with patients at lowest predicted risk receiving less toxic or intensive therapy and being considered for earlier hospital discharge. Encouragingly, this effect was seen in both models, suggesting the ‘basic’ version of the model could be used if measurement of CSF QCC and opening pressure is not possible.

The models demonstrated consistent discrimination across different study sites but demonstrated some variation in calibration, likely reflecting site-level differences in baseline mortality risk not accounted for in the model. In Malawi, the model demonstrated systematic overprediction of risk and in Uganda underprediction, indicating that patients had a less-than and greater-than expected mortality at these sites, respectively. Multiple explanations are plausible, including differences in healthcare provision, genetic predisposition to outcome, and other local socio-economic determinants. Recalibration of the models to individual study sites resolved much of this difference and represents a possible way to adapt the models in future, if required. Nevertheless, in the held-out validation data, discrimination and calibration performance of both ‘basic’ and ‘research’ models improved significantly on the most comparable existing cryptococcal meningitis model for people living with HIV^9^ and is consistent with that achieved to predict mortality in COVID-19.^23^ The final models demonstrated higher net benefit than alternative approaches to inform decision making in decision curve analysis, without recalibration. Notably, most of the observed miscalibration was at higher predicted risks (>25%), where the data were sparser. However, since this represents a risk range above the likely threshold probability for most stratified interventions, this is unlikely to have a significant impact on clinical utility.

Our analyses demonstrated clinically plausible predictor-outcome associations for all included variables in the models and explored non-linear associations of continuous variables with mortality. GCS score <15 and elevated CSF fungal burden have been found to be associated with increased mortality in previous studies from multiple settings.^8,9,12–15,20,21,33^ Notably, CSF fungal burden at baseline, measured using quantitative cultures, had a non-linear association with mortality risk with markedly increased risk above log104, but little increased risk of mortality at lower levels. Although measurement of CSF fungal burden using quantitative cultures is not a routine laboratory procedure, it may be possible in future to utilise results from point-of-care semi-quantitative cryptococcal antigen tests or other approaches to quantify organism load such as qPCR.^34^ We found that ECOG performance status, reflecting functional status, was the most discriminating single predictor for mortality. Since it is easy to measure at presentation, there is a strong rationale for considering this both in research studies and programmatically. Raised intracranial pressure (ICP) is well-known to contribute to morbidity and mortality in HIV-associated cryptococcal meningitis^9,35,36^ and is common on presentation with 580/1488 (39.0%) in the current study having ICP >25cm H2O. The risk associated with 2-week mortality increased modestly with increasing OPs in our analysis. Since raised ICP was managed through protocolised therapeutic lumbar puncture in our included trials, the effect of unmanaged raised ICP on mortality was likely attenuated, as has been hypothesised in previous analyses.^8^ Thus, the association observed in our models should be considered to reflect ICP with protocolised management. Anaemia and high blood neutrophil count were also associated with increased 2-week mortality in our models, as observed for other infectious diseases, and likely reflecting severity of systemic illness.^8^ In our re-trained models for 10-week mortality, age, weight, and seizures on admission were included as additional variables in the ‘basic’ model, while age and CSF cell count were substituted for CSF OP in the ‘research’ model. We hypothesise that most 2-week mortality risk likely relates to cryptococcal meningitis-related pathology, whereas at 10 weeks a larger proportion of the observed risk may be attributable to the underlying HIV and co-morbidities. Since 10-week mortality is significantly higher than 2-week mortality, there were also a larger number of events for the 10-week outcome, which may have led to the inclusion of more variables and slightly better calibration observed for these models.

Though previous studies have sought to compare performance of statistical and machine learning prediction models, most comparisons were classified at high risk of bias in a recent systematic review, due to suboptimal statistical and/or machine learning methodology.^37^ We found that performance of the machine learning model in the validation dataset demonstrated no improvement in performance over the logistic regression model. Our findings suggest that, in lower dimensional datasets, traditional regression approaches may offer equivalent performance whilst remaining computationally less intensive and methodologically more transparent. Notably, our observed predictor-outcome associations from the XGBoost approach largely mirrored the associations found in the regression model.

Strengths of our study include our use of best practices for statistical and machine learning prediction modelling development and validation, including TRIPOD standard reporting, using multiple imputation to deal with missing data, and retaining continuous variables without arbitrary categorisation to avoid loss of information, whilst also accounting for non-linear associations. We also used the largest dataset to date to develop and validate a model for cryptococcal meningitis and mitigated the risk of overfitting our models by defining candidate predictors *a prior*i, in line with best practice sample size guidance.^38^ Furthermore, our pooled dataset included 8 countries; we harnessed this to explore generalisability and geographic heterogeneity in model performance through our internal-external cross-validation approach. Finally, we explored heterogeneity of treatment effect by predicted risk, in line with the PATH statement.^31^

Our study also has limitations. First, our models were developed and validated using data from sub-Saharan Africa, where the greatest burden of disease is; evaluation in other world regions is required to further explore generalisability. While developing the model on RCT data was a strength in terms of data quality, real-world outcomes for cryptococcal meningitis may be inferior to that observed in the trials, due to factors such as trial exclusion criteria and improved standards of care in clinical trials.^39^ Further validation in programmatic cohorts is therefore also required. Whilst our treatment effect analysis provides early proof-of-concept evidence for treatment stratification in cryptococcal meningitis, caution is required due to the exploratory nature of the analysis. While the models were not trained to predict differential treatment effects, their discrimination and calibration to predict 2-week mortality is likely optimistic across participants in the development dataset. Future studies are required to further evaluate differential treatment effects when stratified by our models, including RCTs incorporating approaches to treatment stratification.

In summary, we present prognostic models for 2- and 10-week mortality in HIV-associated cryptococcal meningitis and demonstrate consistent performance across sub-Saharan African settings. The models use commonly available predictors and will be made freely available to direct future treatment stratification approaches in clinical trials (prototype: https://rishi-k-gupta.shinyapps.io/periskope-cm/).

## Acknowledgements

We would like to acknowledge all site staff and participants contributing to the primary randomised-controlled trials.

## Author contributions

SFM, TSH, JNJ and RKG conceived the study. THAS, SFM, TSH, JNJ and RKG wrote the analysis plan. SFM curated the pooled dataset. THAS and RKG performed the analysis. THAS, SFM and RKG wrote the first draft of the manuscript, with input from TSH and JNJ. RKG built the Rshiny app and WSL developed the PERISKOPE website. All contributing authors reviewed and approved the manuscript prior to submission.

## Data availability statement

The individual-level data for the trials included in this pooled analysis will be made available to investigators upon reasonable request to the corresponding authors of the original trials.

