## Supplementary Appendix for "Personalised risk prediction tools for cryptococcal meningitis mortality to guide treatment stratification; a pooled analysis of two randomised-controlled trials"

### Supplementary Methods

#### *Sample size estimation*

A total of 243/1488 participants (16.3%) reached the primary outcome (2-week mortality). There are no currently widely used tools to predict mortality in HIV-associated cryptococcal meningitis. In other infectious diseases, the 4C mortality score is a widely used prediction tool to predict in-hospital mortality in COVID-19[1] and achieved a C-statistic of 0.8 in a recent individual participant data meta-analysis.[2] Based on this target C-statistic of 0.8, we estimated that 28 parameters could be considered for inclusion in the final model. [3]

#### *XGBoost modelling*

XGBoost is an ensemble, gradient boosted decision tree method where a series of decision trees is constructed, with each tree seeking to sequentially minimise errors from preceding trees.

A range of XGBoost hyper-parameters were evaluated in the development dataset using a grid search, where default parameters were varied (Supplementary Table 2). We identified the best combination of hyper-parameters using 10-fold cross-validation in the development set, defined as the highest C-statistic.

A common critique of machine learning methods such as XGBoost is that they represent “black box” methods, where the plausibility of predictor-outcome associations cannot readily be evaluated. [4] To mitigate this criticism, we explored predictor-outcome associations in the final XGBoost model by using the model object to make predictions across a range of values for each predictor in turn, while fixing all other variables to the median (for continuous variables) or modal (for factor variables) values. We then plotted these associations visually for each variable. Furthermore, since machine learning models such as decision trees can include any number of interactions without prior specification, we also examined the presence of two-way interactions by varying 2 variables at a time. The first of these predictors was included as the x-axis, while the second was coloured as a factor (using the midpoint of quartiles from the observed data for continuous predictors). This enabled construction of a matrix of plots visualising XGBoost predictor-outcome associations, while examining for all possible two-way interactions.

The XGBoost model was trained and validated using stacked development and validation datasets including the 10 multiply imputed sets, respectively.

#### *Code Sharing*

All analytical code used to generate the results in this manuscript will be made publicly available in a GitHub repository, including all R packages used with versions.

#### *References*

1. Knight SR, Ho A, Pius R, Buchan I, Carson G, Drake TM, et al. Risk stratification of patients admitted to hospital with covid-19 using the ISARIC WHO Clinical Characterisation Protocol: Development and validation of the 4C Mortality Score. BMJ. 2020 Sep 9;370:22.

2. De Jong VMT, Rousset RZ, Antonio-Villa NE, Buenen AG, Van Calster B, Bello-Chavolla OY, et al. Clinical prediction models for mortality in patients with covid-19: external validation and individual participant data meta-analysis. BMJ. 2022 Jul 12;378:39.

3. Riley RD, Ensor J, Snell KIE, Harrell FE, Martin GP, Reitsma JB, et al. Calculating the sample size required for developing a clinical prediction model. BMJ. 2020 Mar 18;368.

4. Murdoch WJ, Singh C, Kumbier K, Abbasi-Asl R, Yu B. Definitions, methods, and applications in interpretable machine learning. Proc Natl Acad Sci U S A. 2019 Oct 29;116(44):22071–80.

### Supplementary Results

#### *Supplementary Table 1: Candidate predictors for multivariable models*

Predictors were rationalized *a priori* in accordance with the number of parameters considered for inclusion in the sample size calculation.

| Predictor | Model Inclusion | Variable type | Parameters included (including spline transformations) |
|---|---|---|---|
| Age | Basic and Research | Continuous | 2 |
| Sex | Basic and Research | Binary | 1 |
| Weight | Basic and Research | Continuous | 2 |
| Seizures | Basic and Research | Binary | 1 |
| Glasgow Coma Score (GCS) | Basic and Research | Factor (15, 10-14, <10) | 2 |
| Eastern Cooperative Group (ECOG) performance status | Basic and Research | Factor (0, 1, 2, 3, 4) | 4 |
| Neutrophil cell count | Basic and Research | Continuous | 2 |
| Haemoglobin | Basic and Research | Continuous | 2 |
| CD4 count | Research only | Continuous | 2 |
| Cerebrospinal fluid (CSF) opening pressure | Research only | Continuous | 2 |
| CSF white cell count (WCC) | Research only | Continuous | 2 |
| Fungal burden in CSF | Research only | Continuous (log base 10 transformed) | 2 |
| Treatment arm | Basic and Research | Factor<br>(Liposomal-Amphotericin-B Ambition regimen and the 1-week Amphotericin-B + Flucytosine arms from both ACTA and Ambition-cm trials,<br>1 week Amphotericin-B + Fluconazole,<br>2 weeks Amphotericin-B + Flucytosine,<br>2 weeks Amphotericin-B + Fluconazole,<br>Flucytosine + Fluconazole oral regimen) | 4 |

***Supplementary Table 2: Hyper-parameters for XGBoost***

| Parameter | Default values | Grid search values | Final selected value |
|---|---|---|---|
| nrounds | 100 | 50, 100, 150 | 50 |
| eta | 6 | 4, 6, 8 | 0.1 |
| max_depth | 0.3 | 0.1, 0.3, 0.5 | 4 |
| gamma | 0 | 0, 1 | 0 |
| colsample_bytree | 1 | 0.8, 1 | 0.8 |
| min_child_weight | 1 | 1, 2, 3 | 2 |
| subsample | 1 | 0.8, 1 | 0.8 |

***Supplementary Figure 1: Schematic showing analysis pipeline.***

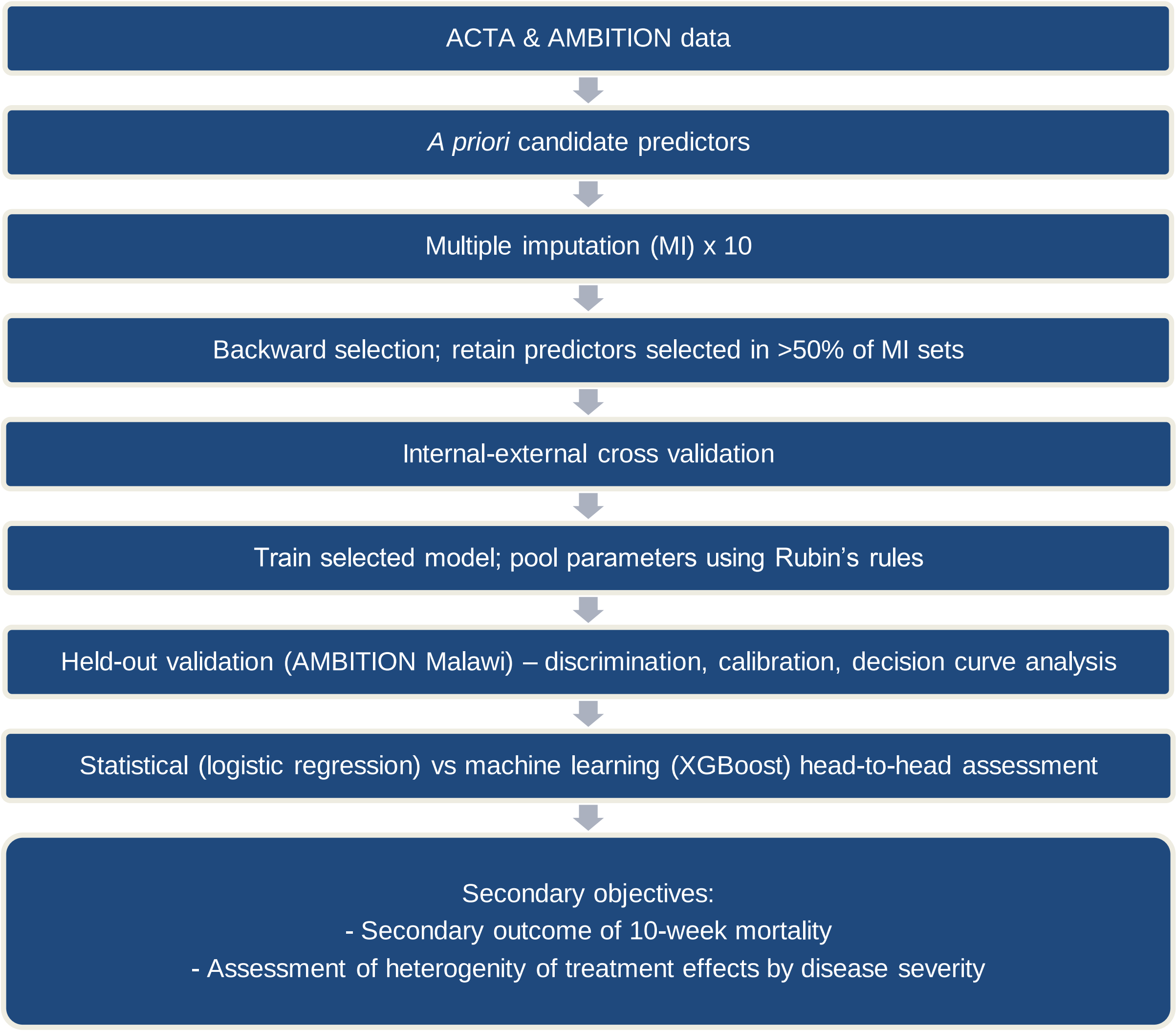

*Supplementary Figure 2: Participant Flow Diagram*

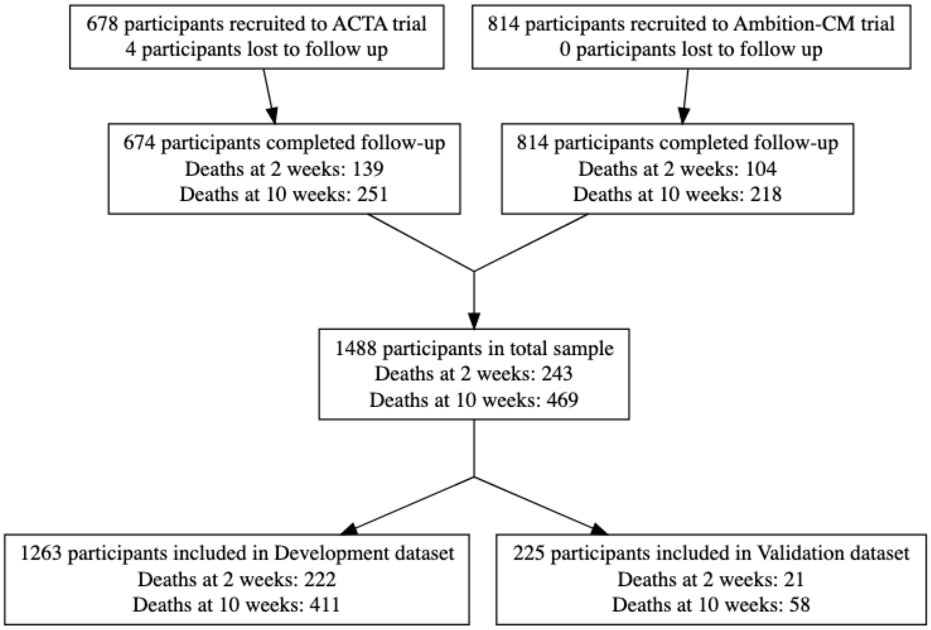

***Supplementary Table 3: Participant characteristics split by development and validation datasets***

ECOG = Eastern Cooperative Oncology Group performance status; GCS = Glasgow Coma Scale; CSF = cerebrospinal fluid.

| **Characteristic** | **Overall**, N = 1,488[1] | **Development**, N = 1,263[1] | **Validation**, N = 225[1] |
|---|---|---|---|
| Mortality at 2 weeks | | | |
| Alive | 1,245 (84%) | 1,041 (82%) | 204 (91%) |
| Died | 243 (16%) | 222 (18%) | 21 (9.3%) |
| Age (years) | 37 (32, 43) | 37 (32, 43) | 38 (32, 44) |
| Sex | | | |
| Female | 612 (41%) | 526 (42%) | 86 (38%) |
| Male | 876 (59%) | 737 (58%) | 139 (62%) |
| Weight (kg) | 52 (47, 60) | 52 (47, 60) | 53 (46, 60) |
| Missing | 15 | 15 | 0 |
| Seizures | 204 (14%) | 176 (14%) | 28 (12%) |
| Missing | 4 | 4 | 0 |
| GCS score | | | |
| 15 | 1,095 (74%) | 929 (74%) | 166 (74%) |
| 11-14 | 327 (22%) | 282 (22%) | 45 (20%) |
| <=10 | 66 (4.4%) | 52 (4.1%) | 14 (6.2%) |
| ECOG performance status | | | |
| Normal | 64 (4.3%) | 57 (4.5%) | 7 (3.1%) |
| Restricted activity | 256 (17%) | 217 (17%) | 39 (17%) |
| Ambulatory | 339 (23%) | 299 (24%) | 40 (18%) |
| Limited self-care | 512 (34%) | 425 (34%) | 87 (39%) |
| Bedbound | 316 (21%) | 264 (21%) | 52 (23%) |
| Missing | 1 | 1 | 0 |
| White cell count (x10^9/L) | 4.20 (3.10, 5.60) | 4.20 (3.10, 5.69) | 4.30 (3.10, 5.50) |
| Missing | 12 | 12 | 0 |
| Neutrophil count (x10^9/L) | 2.50 (1.66, 3.80) | 2.50 (1.69, 3.86) | 2.29 (1.64, 3.33) |
| Missing | 31 | 30 | 1 |
| Haemoglobin (g/L) | 110 (96, 126) | 110 (96, 126) | 112 (97, 125) |
| Missing | 10 | 10 | 0 |

| Characteristic | **Overall**, N = 1,488[1] | **Development**, N = 1,263[1] | **Validation**, N = 225[1] |
|---|---|---|---|
| CD4 count (x10^6/L) | 27 (10, 62) | 26 (10, 60) | 35 (12, 67) |
| Missing | 88 | 73 | 15 |
| CSF opening pressure (cmH20) | 22 (13, 33) | 22 (14, 35) | 20 (12, 30) |
| Missing | 54 | 54 | 0 |
| CSF cell count (WBC per mm3) | 4 (1, 37) | 4 (1, 32) | 17 (3, 68) |
| Missing | 59 | 46 | 13 |
| CSF Glucose (mmol/L) | 2.10 (1.21, 3.00) | 2.10 (1.20, 3.00) | 2.10 (1.38, 2.80) |
| Missing | 120 | 119 | 1 |
| CSF Protein (mg/mL) | 0.93 (0.50, 1.54) | 0.90 (0.48, 1.49) | 1.06 (0.54, 1.70) |
| Missing | 127 | 122 | 5 |
| log(CSF quantitative culture) | 4.79 (3.08, 5.66) | 4.81 (3.19, 5.67) | 4.61 (2.73, 5.57) |
| Missing | 40 | 40 | 0 |

[1]n (%); Median (IQR)

***Supplementary Figure 3: Multivariable associations between selected predictors and outcome in basic primary model***

Continuous variables were modeled using restricted cubic splines. The final model parameters are pooled across multiply imputed datasets (total sample size for model development = 1,263 participants). For continuous variables, black lines represent point estimates and grey shaded regions represent 95% confidence intervals. For categorical variables, black dots represent point estimates and black lines represent 95% confidence intervals. ECOG = Eastern Cooperative Oncology Group performance status; GCS = Glasgow Coma Scale.

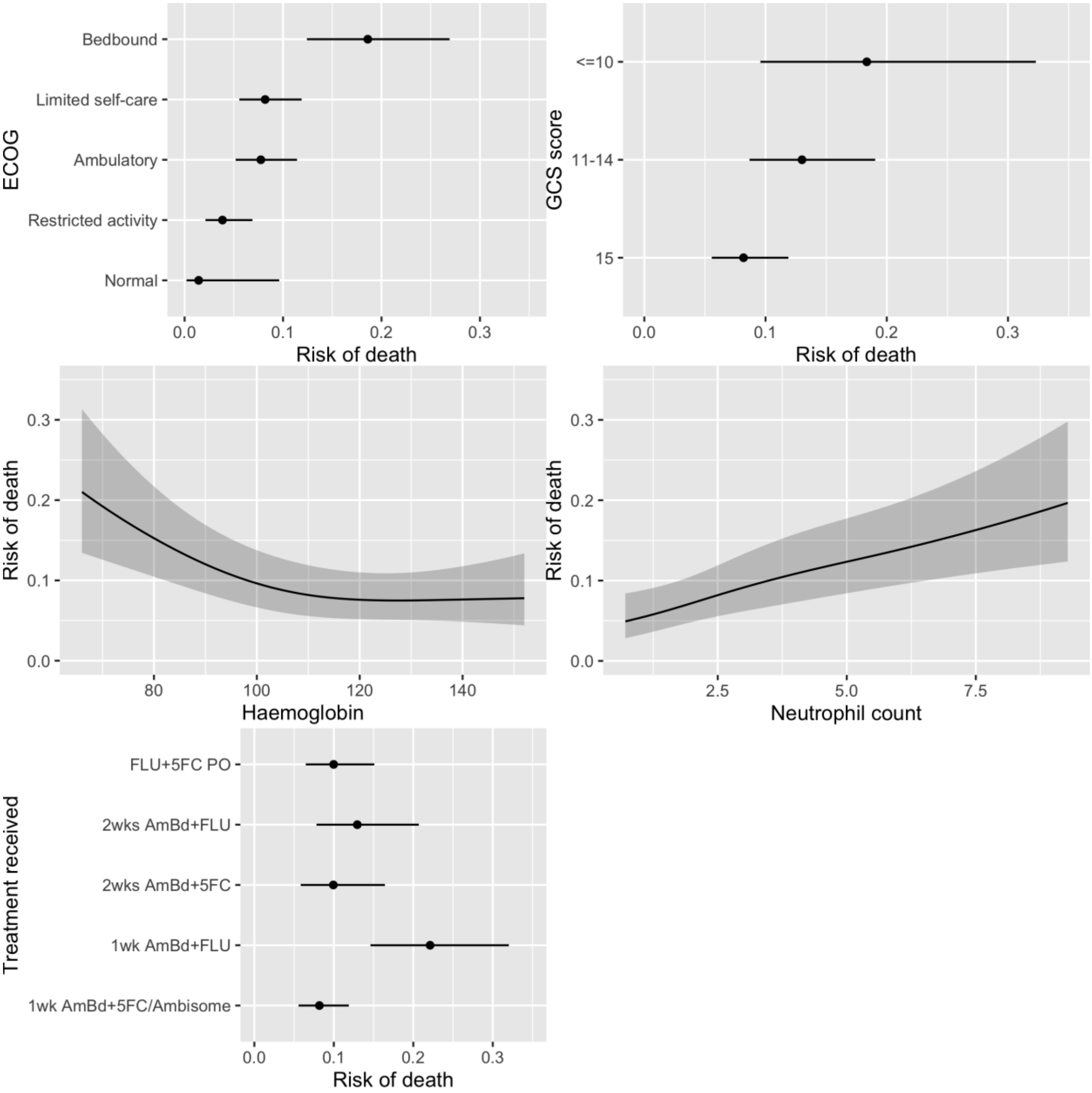

***Supplementary Table 4 and 5: Pooled model parameters for basic and research models***

Relationships between independent predictors and outcome are represented as odds ratios. ECOG = Eastern Cooperative Oncology Group performance status; CSF = cerebrospinal fluid; QCC = quantitative cryptococcal culture; AmBd = Amphotericin B deoxycholate; 5FC = Flucytosine; Flu = Fluconazole.

*Supplementary Table 4: Basic Model*

| Variable | Estimate | 95% Confidence Int. |
|---|---|---|
| Intercept | 0.13 | 0.01 to 1.45 |
| Glasgow Coma Score: 11-14 | 1.68 | 1.13 to 2.49 |
| Glasgow Coma Score: ≤10 | 2.52 | 1.27 to 5.02 |
| ECOG: Restricted activity | 2.76 | 0.35 to 21.69 |
| ECOG: Ambulatory | 5.79 | 0.77 to 43.59 |
| ECOG: Limited self-care | 6.15 | 0.82 to 45.99 |
| ECOG: Bedbound | 15.77 | 2.08 to 119.57 |
| Treatment: 1wk AmBd+Flu | 3.19 | 1.92 to 5.28 |
| Treatment: 2wk AmBd+5FC | 1.24 | 0.72 to 2.14 |
| Treatment: 2wk AmBd+Flu | 1.67 | 0.97 to 2.87 |
| Treatment: 5FC+Flu | 1.24 | 0.80 to 1.93 |
| Neutrophils | 1.38 | 1.05 to 1.80 |
| Neutrophils (spline 1) | 0.81 | 0.57 to 1.16 |
| Haemoglobin | 0.97 | 0.96 to 0.99 |
| Haemoglobin (spline 1) | 1.02 | 1.00 to 1.04 |

*Supplementary Table 5: Research Model*

| Variable | Estimate | 95% Confidence Int. |
|---|---|---|
| Intercept | 0.18 | 0.02 to 2.29 |
| Glasgow Coma Score: 11-14 | 1.73 | 1.14 to 2.61 |
| Glasgow Coma Score: ≤10 | 3.23 | 1.57 to 6.63 |
| ECOG: Restricted activity | 2.53 | 0.32 to 20.18 |
| ECOG: Ambulatory | 5.40 | 0.71 to 41.05 |
| ECOG: Limited self-care | 5.34 | 0.71 to 40.33 |
| ECOG: Bedbound | 13.45 | 1.75 to 103.32 |
| Treatment: 1wk AmBd+Flu | 2.86 | 1.68 to 4.85 |
| Treatment: 2wk AmBd+5FC | 1.19 | 0.68 to 2.10 |
| Treatment: 2wk AmBd+Flu | 1.53 | 0.86 to 2.72 |
| Treatment: 5FC+Flu PO | 1.20 | 0.76 to 1.89 |
| Neutrophils | 1.30 | 0.99 to 1.72 |
| Neutrophils (spline 1) | 0.89 | 0.62 to 1.28 |
| Haemoglobin | 0.97 | 0.95 to 0.98 |
| Haemoglobin (spline 1) | 1.02 | 1.00 to 1.04 |
| CSF Opening Pressure | 1.00 | 0.97 to 1.03 |
| CSF Opening Pressure (spline 1) | 1.02 | 0.98 to 1.05 |
| CSF QCC (log) | 0.77 | 0.62 to 0.96 |
| CSF QCC (log) (spline 1) | 1.65 | 1.36 to 2.01 |

***Supplementary Figure 4: Pooled calibration plots across multiply imputed development datasets, both before and after recalibration of intercepts to country of origin.***

a) basic model; b) research model; c) basic model post-recalibration; d) research model post-recalibration. Plots in red are before recalibration and plots in blue are post-recalibration. Calibration is shown using a loess smoother. Rug plots, shown on the x-axis, plot the distribution of predicted risk.

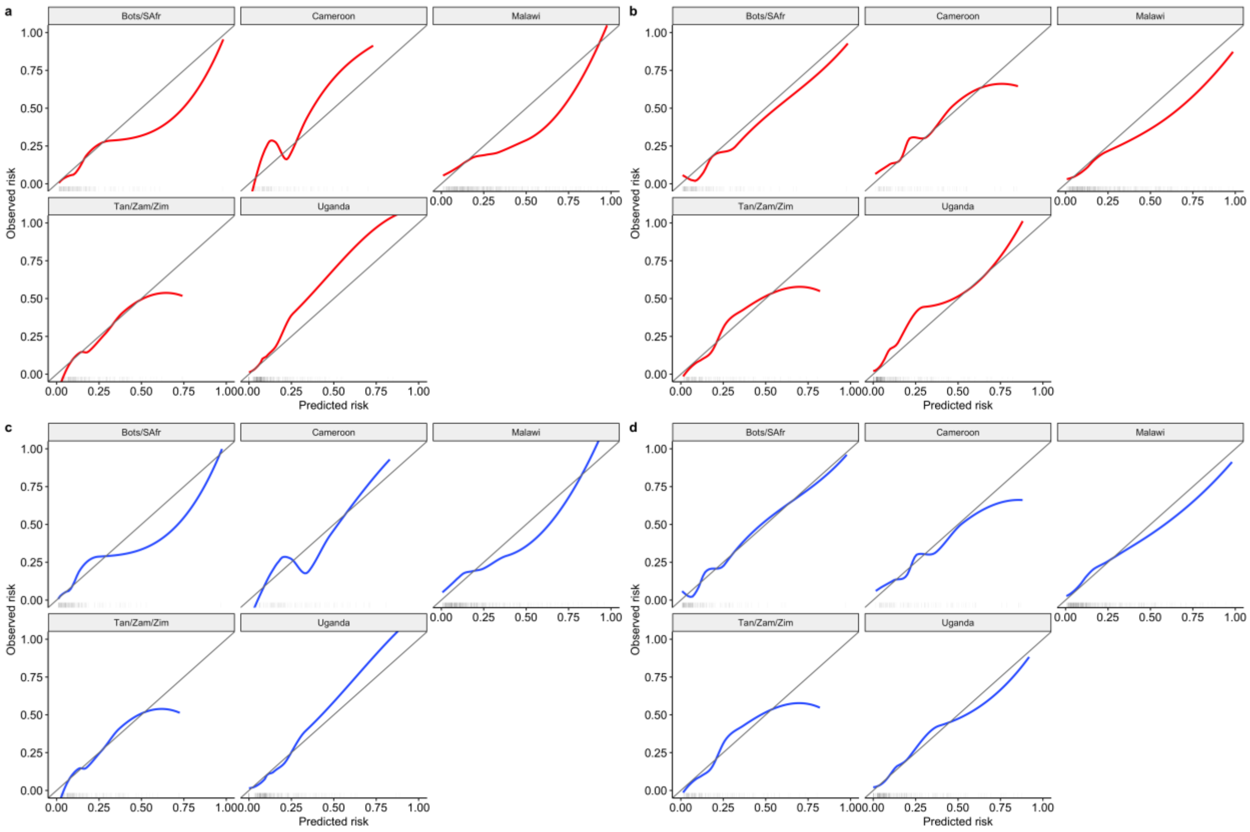

***Supplementary Figure 5: Calibration of Zhao et al. model***

Calibration is shown using a loess-smoother across multiply imputed datasets. The original variable coefficients were extracted from Zhao et al's model, a model intercept was derived from our validation data, and a regression model constructed to allow the calculation of predicted risk. The rug plot indicates the distribution of predicted risk.

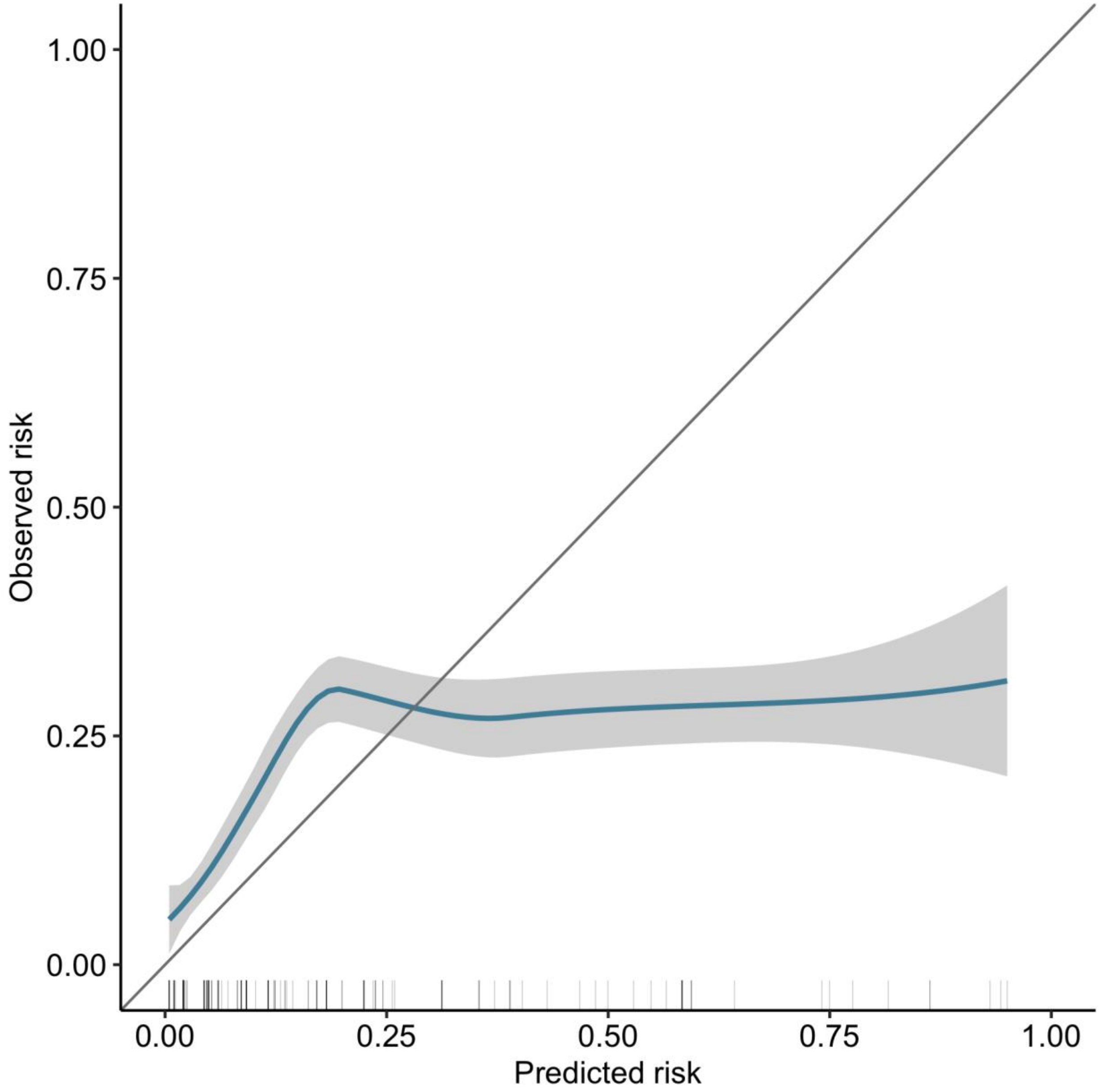

***Supplementary Figure 6: Decision Curve Analysis in held-out validation data***

Net benefit is shown for each candidate model with loess smoothing, compared to ‘treat all’ and ‘treat none’ approaches. As for supplementary figure 5, predictions from a regression model derived from Zhao model coefficients were used to avoid over-optimistic estimates of net benefit.

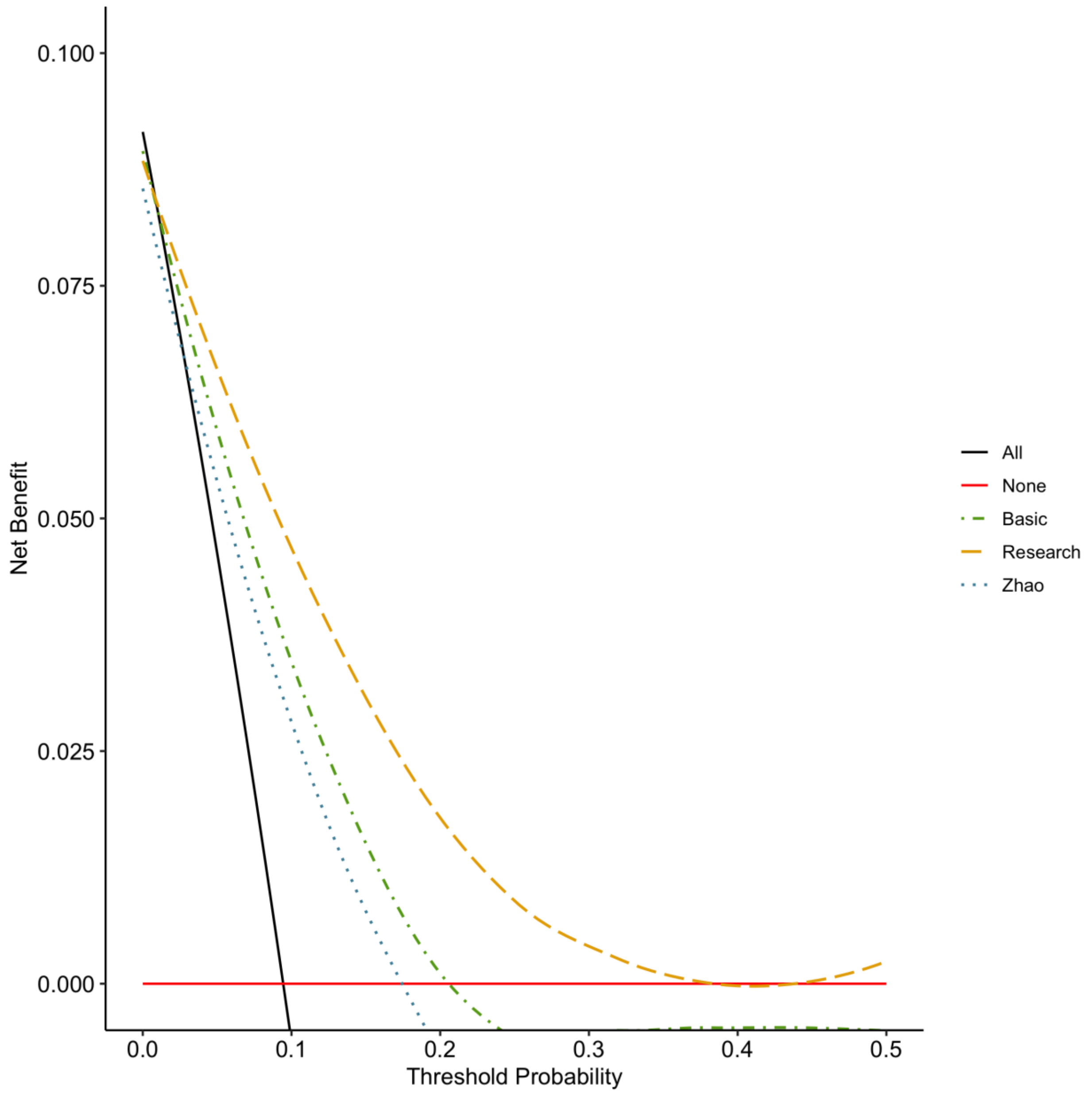

***Supplementary Table 6: Performance of single predictors included in main models in held-out validation data***

Individual factors making up both models were assessed for discriminatory ability against held-out validation data from Ambition trial in Malawi. Results sorted by C-statistic. Treatment arm was not included as all participants in the validation cohort received the same treatment factor level (1 week Amphotericin B + flucytosine or high-dose liposomal Amphotericin B.). ECOG = Eastern Cooperative Oncology Group performance status; CSF = cerebrospinal fluid.

| Predictor | AUROC[1,2] |
|---|---|
| ECOG Performance Status | 0.78 (0.71 - 0.85) |
| CSF Quantitative Culture (log cfu/ml) | 0.72 (0.59 - 0.85) |
| Glasgow Coma Score | 0.68 (0.56 - 0.79) |
| CSF Opening Pressure (cmH20) | 0.67 (0.54 - 0.79) |
| Neutrophils (x10^9/L) | 0.66 (0.53 - 0.79) |
| Haemoglobin (g/L) | 0.55 (0.40 - 0.69) |

[1]AUROC = Area under the receiver operator characteristic curve

[2]Brackets show 95% confidence intervals

***Supplementary Table 7: Performance of single predictors included in main models by country in development data***

Individual predictors making up both models were assessed for discriminatory ability in an internal-external cross validation approach in the development dataset. GCS = Glasgow Coma Scale; ECOG = Eastern Cooperative Oncology Group performance status; CSF = cerebrospinal fluid; Tan = Tanzania; Zam = Zambia; Zim = Zimbabwe; Bots = Botswana; SAfr = South Africa.

| Location | AUROC[1,2] | | | | | | |
|---|---|---|---|---|---|---|---|
| | ECOG | CSF Quantitative Culture | Neutrophils | GCS | CSF Opening Pressure | Haemoglobin | Treatment |
| Uganda | 0.77 | 0.59 | 0.72 | 0.73 | 0.60 | 0.59 | 0.50 |
| Tan/Zam/Zim | 0.67 | 0.64 | 0.60 | 0.58 | 0.59 | 0.69 | 0.65 |
| Malawi | 0.66 | 0.68 | 0.60 | 0.62 | 0.53 | 0.54 | 0.55 |
| Cameroon | 0.64 | 0.61 | 0.63 | 0.57 | 0.63 | 0.54 | 0.62 |
| Bots/SAfr | 0.78 | 0.67 | 0.65 | 0.64 | 0.59 | 0.51 | 0.50 |

[1]AUROC = Area under the receiver operator characteristic curve

[2]C-statistic results coloured from a lighter to darker blue as C-statistic increases

***Supplementary Figure 7: Associations between component variables in machine learning model***

Matrix plot in which each column represents associations with that component variable of the model. Plots shown in grey on the diagonal represent the multivariable relationship between that variable and the mortality outcome. Other plots in the column represent the interaction between the column variable and the labelled row variables. Where the row variable is categorical, these interactions are plotted as separate lines representing the relationship between different levels of the row variable. Where the row variable is continuous, these lines represent quartiles of this variable. Treatment arm 1 through 5 represent 1) the liposomal-Amphotericin-B Ambition regimen and the 1-week Amphotericin-B + Flucytosine arms from both ACTA and Ambition-cm trials, 2) 1 week Amphotericin-B + Fluconazole, 3) 2 weeks Amphotericin-B + Flucytosine, 4) 2 weeks Amphotericin-B + Fluconazole and 5) Flucytosine + Fluconazole oral regimen, respectively.

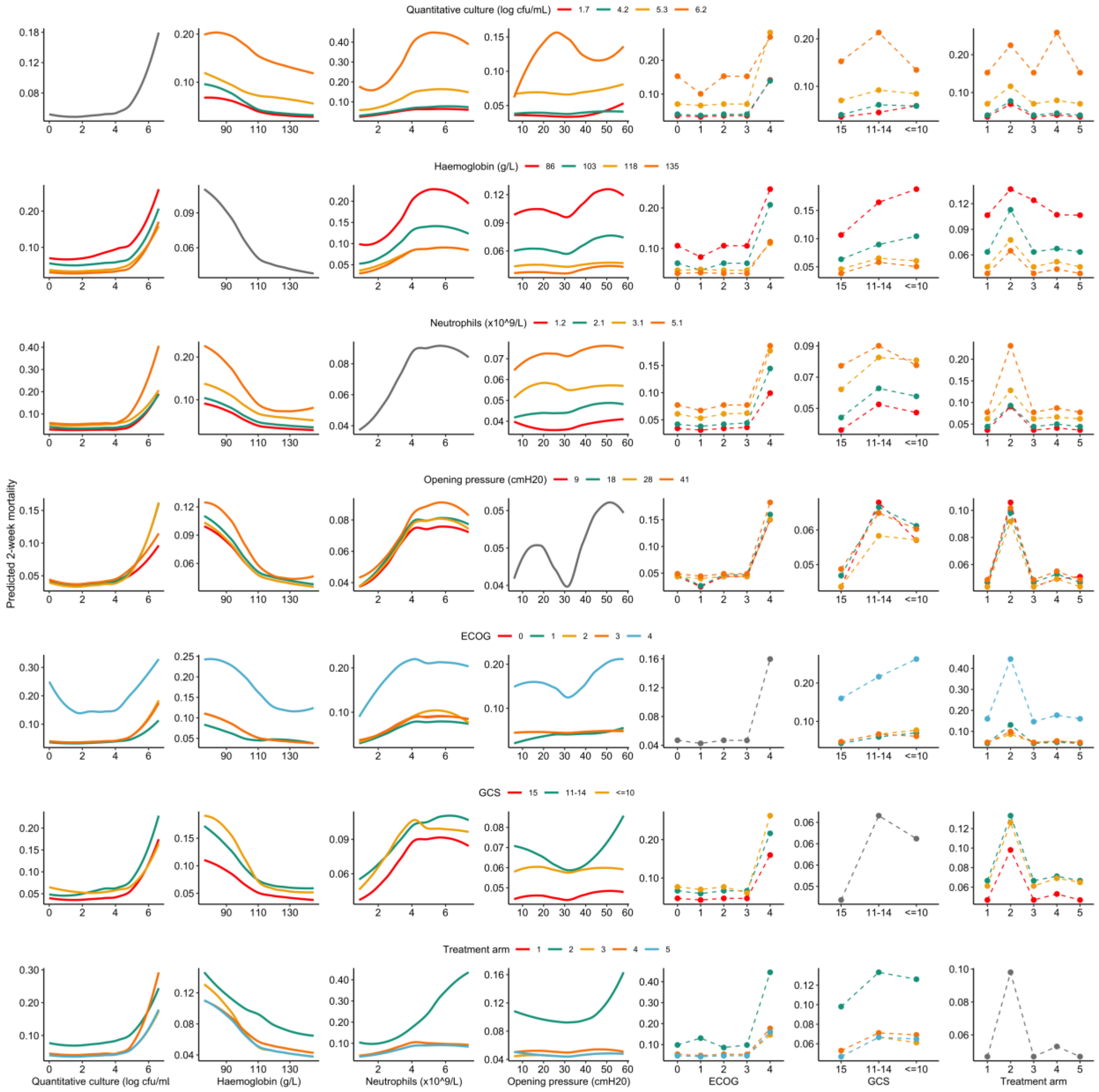

***Supplementary Figure 8: Internal-external cross validation results for the XGBoost Machine Learning model***

Pooled estimates are calculated through random-effects meta-analysis (total sample size = 1,263 participants). Countries with n < 100 participants or x < 20 deaths were amalgamated and grouped by similarity of healthcare environment. Dashed lines indicate lines of perfect calibration in the large (0) and slope (1), respectively. Black squares indicate point estimates; bars indicate 95% confidence intervals; diamonds indicate pooled random-effects meta-analysis estimates. Bots = Botswana; SAfr = South Africa; Tan = Tanzania; Zam = Zambia; Zim = Zimbabwe.

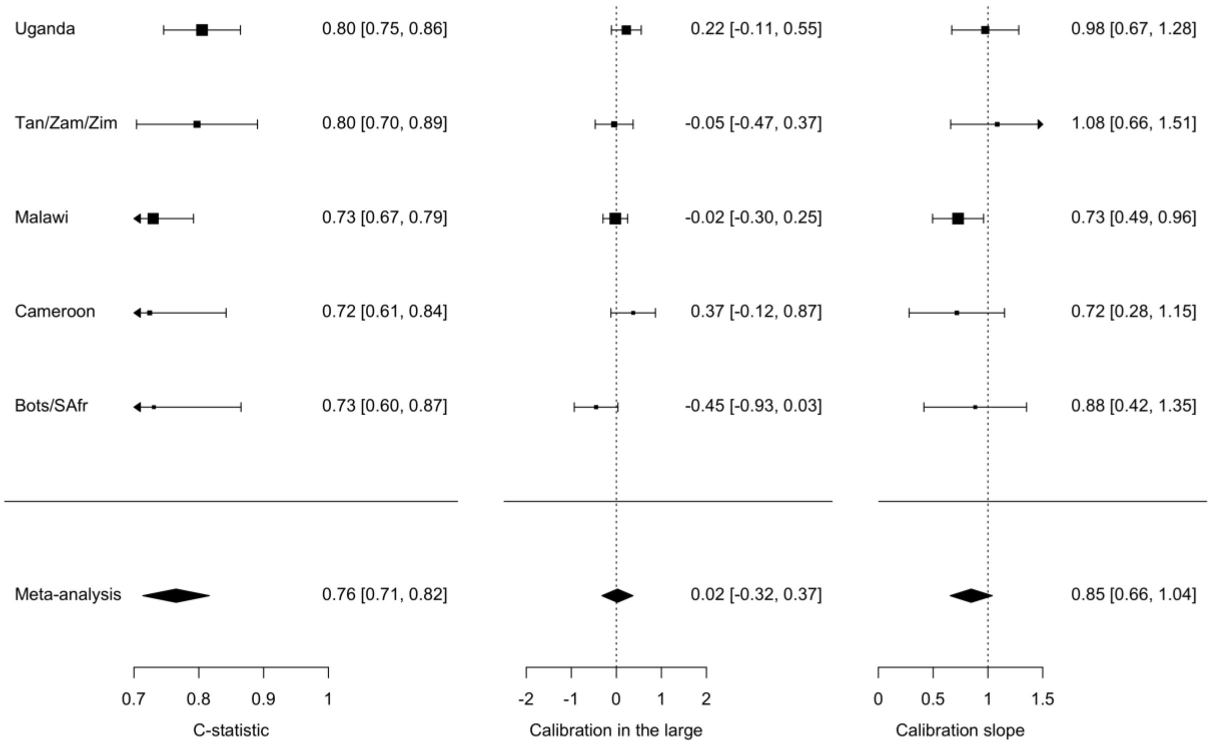

***Supplementary Table 8: Variables selected into retrained 10-week mortality model***

Variable selection was done in each imputed dataset using backward elimination using AIC. Variables retained in >50% of multiply imputed datasets were selected into the model. ECOG = Eastern Cooperative Oncology Group performance status; CSF = cerebrospinal fluid; MI = multiple imputation.

| | **Baseline Model** | **Research Model** |
|---|---|---|
| **Variable** | Number of MI datasets selected (total n=10)[1] | Number of MI datasets selected (total n=10)[1] |
| Age | 10 (100%) | 10 (100%) |
| Sex | 0 (0%) | 0 (0%) |
| Weight | 10 (100%) | 3 (30%) |
| Seizure | 10 (100%) | 0 (0%) |
| GCS | 0 (0%) | 9 (90%) |
| ECOG | 10 (100%) | 10 (100%) |
| Treatment regimen | 10 (100%) | 10 (100%) |
| Neutrophil count | 10 (100%) | 10 (100%) |
| Haemoglobin | 10 (100%) | 10 (100%) |
| CD4 count | | 0 (0%) |
| CSF opening pressure | | 0 (0%) |
| CSF cell count | | 7 (70%) |
| CSF Quantitative Culture (log) | | 10 (100%) |

[1]n (%)

***Supplementary Figure 9: Multivariable model associations in predictors selected in retrained 10-week model***

Supplementary Figure 9a shows the associations in the retrained basic model and is shown first, with 1b showing the associations in the retrained research model. Continuous variables were modeled using restricted cubic splines. The final model parameters are pooled across multiply imputed datasets (total sample size for model development = 1,263 participants). For continuous variables, black lines represent point estimates and grey shaded regions represent 95% confidence intervals. For categorical variables, black dots represent point estimates and black lines represent 95% confidence intervals. ECOG = Eastern Cooperative Oncology Group performance status; GCS = Glasgow Coma Scale; CSF = cerebrospinal fluid.

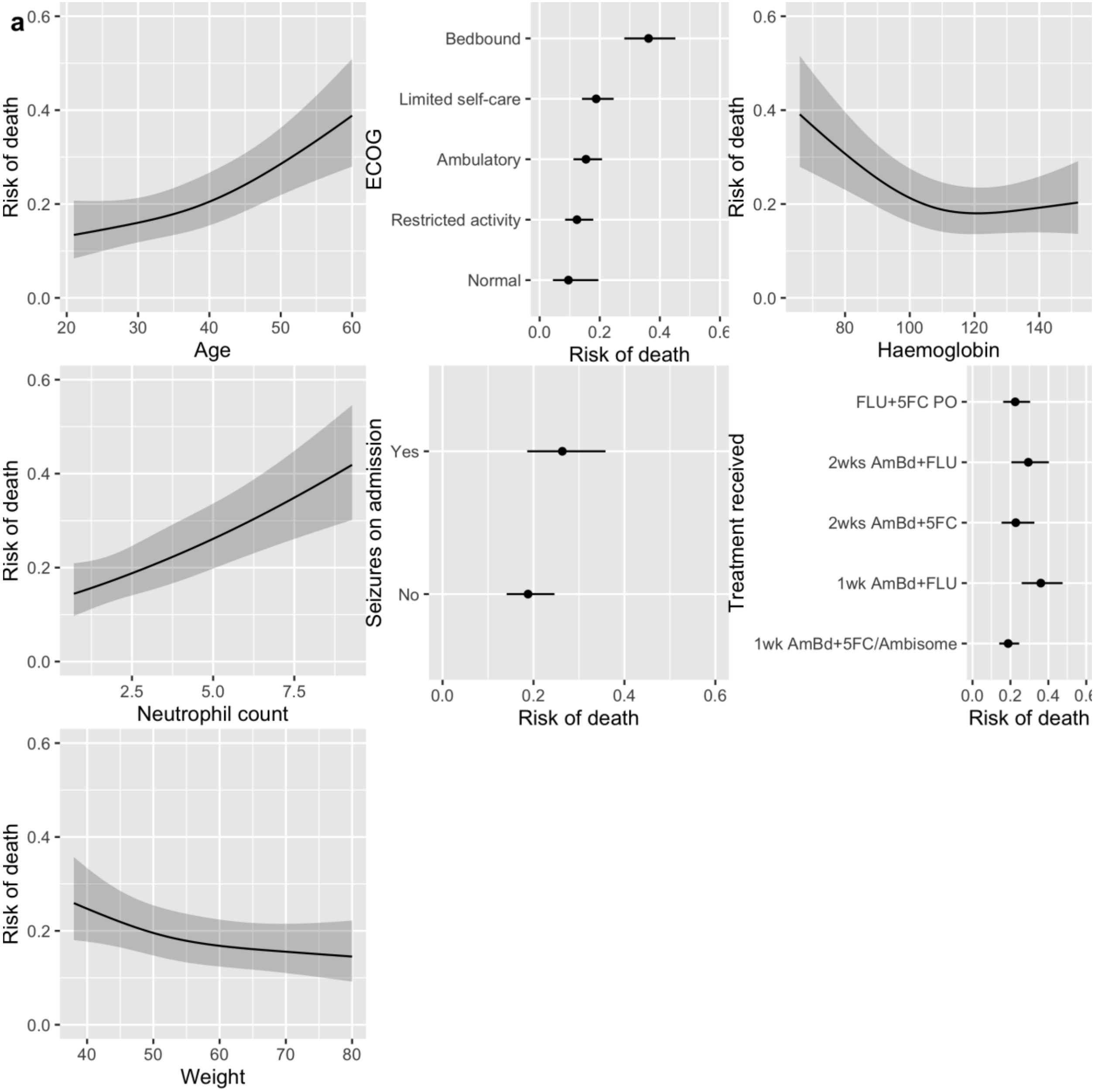

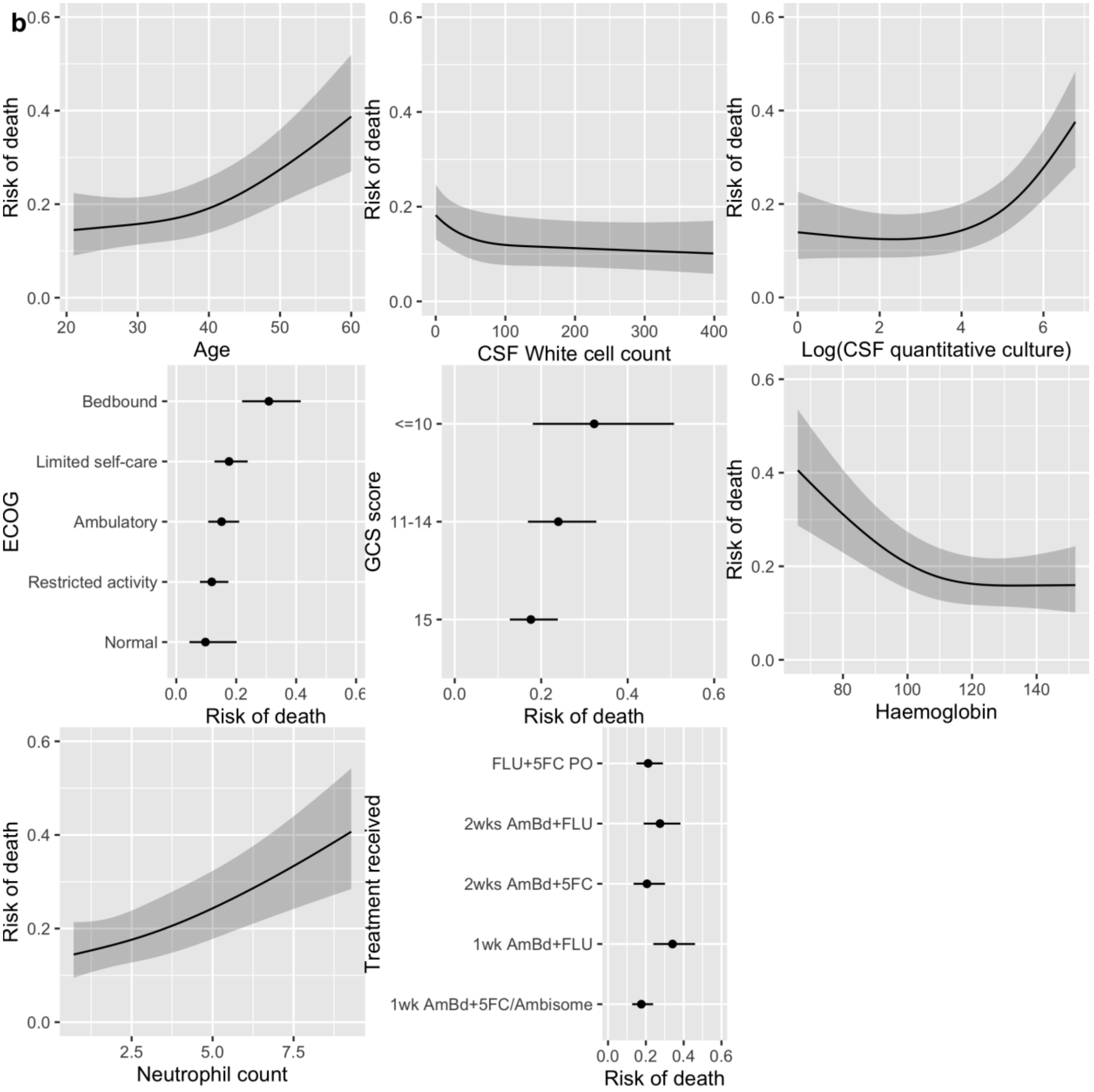
b
Risk of death
Age
CSF White cell count
Log(CSF quantitative culture)
ECOG
Bedbound
Limited self-care
Ambulatory
Restricted activity
Normal
GCS score
<=10
11-14
15
Haemoglobin
Neutrophil count
Treatment received
FLU+5FC PO
2wks AmBd+FLU
2wks AmBd+5FC
1wk AmBd+FLU
1wk AmBd+5FC/Ambisome

***Supplementary Tables 9 and 10: Pooled model parameters for (a) basic and (b) research 10-week model***

Relationships between independent predictors and outcome are represented as odds ratios. ECOG = Eastern Cooperative Oncology Group performance status; CSF = cerebrospinal fluid; QCC = quantitative cryptococcal culture; AmBd = Amphotericin B deoxycholate; 5FC = Flucytosine; Flu = Fluconazole.

*(a)*

| Variable | Estimate | 95% Confidence Int. |
|---|---|---|
| Intercept | 2.25 | 0.25 to 20.08 |
| Age | 1.02 | 0.99 to 1.06 |
| Age (spline 1) | 1.02 | 0.98 to 1.06 |
| Weight | 0.97 | 0.94 to 1.00 |
| Weight (spline 1) | 1.02 | 0.98 to 1.06 |
| Seizure: Yes | 1.55 | 1.07 to 2.22 |
| ECOG: Restricted activity | 1.34 | 0.55 to 3.24 |
| ECOG: Ambulatory | 1.72 | 0.73 to 4.06 |
| ECOG: Limited self-care | 2.19 | 0.94 to 5.09 |
| ECOG: Bedbound | 5.37 | 2.28 to 12.66 |
| Treatment: 1wk AmBd+Flu | 2.43 | 1.56 to 3.80 |
| Treatment: 2wk AmBd+5FC | 1.28 | 0.82 to 2.00 |
| Treatment: 2wk AmBd+Flu | 1.80 | 1.15 to 2.82 |
| Treatment: 5FC+Flu | 1.26 | 0.88 to 1.78 |
| Neutrophils | 1.19 | 0.97 to 1.48 |
| Neutrophils (spline 1) | 0.99 | 0.74 to 1.32 |
| Haemoglobin | 0.97 | 0.96 to 0.99 |
| Haemoglobin (spline 1) | 1.02 | 1.01 to 1.04 |

*(b)*

| Variable | Estimate | 95% Confidence Int. |
|---|---|---|
| Intercept | 0.86 | 0.13 to 5.58 |
| Age | 1.01 | 0.98 to 1.05 |
| Age (spline 1) | 1.03 | 0.99 to 1.08 |
| Glasgow Coma Score: 11-14 | 1.48 | 1.04 to 2.08 |
| Glasgow Coma Score: ≤10 | 2.23 | 1.10 to 4.50 |
| ECOG: Restricted activity | 1.25 | 0.51 to 3.05 |
| ECOG: Ambulatory | 1.65 | 0.69 to 3.94 |
| ECOG: Limited self-care | 1.98 | 0.84 to 4.68 |
| ECOG: Bedbound | 4.15 | 1.69 to 10.18 |
| Treatment: 1wk AmBd+Flu | 2.42 | 1.53 to 3.83 |
| Treatment: 2wk AmBd+5FC | 1.21 | 0.76 to 1.93 |
| Treatment: 2wk AmBd+Flu | 1.77 | 1.12 to 2.81 |
| Treatment: 5FC+Flu PO | 1.26 | 0.87 to 1.81 |
| Neutrophils | 1.14 | 0.92 to 1.41 |
| Neutrophils (spline 1) | 1.06 | 0.79 to 1.42 |
| Haemoglobin | 0.97 | 0.96 to 0.98 |
| Haemoglobin (spline 1) | 1.02 | 1.00 to 1.04 |
| CSF White Cell Count | 0.99 | 0.98 to 1.00 |
| CSF White Cell Count (spline 1) | 1.12 | 0.99 to 1.26 |
| CSF QCC (log) | 0.93 | 0.78 to 1.11 |
| CSF QCC (log) (spline 1) | 1.33 | 1.14 to 1.56 |

***Supplementary Figure 10: Calibration plots of retrained 10-week mortality model in held-out validation data***

(a) Basic Treatment Model; (b) Research Treatment Model

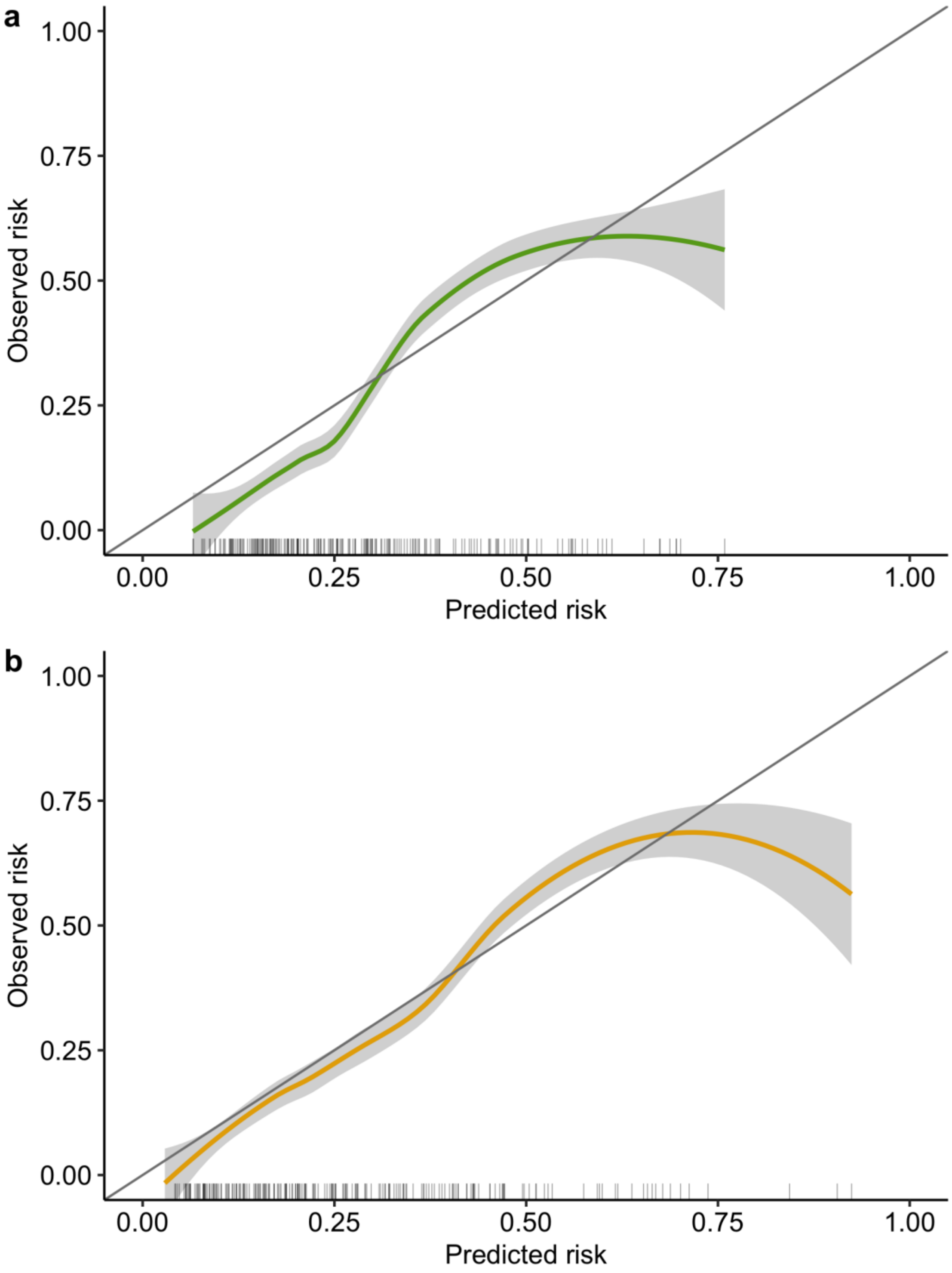

***Supplementary Figure 11: Distribution of predicted mortality risk by risk tercile.***

Boxplots and density plots showing the distribution of predicted 2-week mortality across the total MI cohort. Boxplots A and C show the distribution of risk by tercile, as derived from the Basic Model (A) and Research Model (C). Horizontal dashed lines represent the threshold of predicted mortality delineating each tercile. Tercile thresholds in the basic model predictions were 0.081 and 0.181 for Low/Medium Risk and Medium/High Risk respectively. Tercile thresholds in the research model were 0.064 and 0.171 respectively. Density plots B and D show the distribution of predicted risk across the total MI cohort, derived from the Basic Model (B) and Research Model (D). Vertical dashed lines delineate the tercile-derived thresholds of risk from (A) and (C), respectively. For the Basic Model, the median (IQR) mortality risk prediction was 0.117 (0.068 to 0.234) and the modal prediction was 0.076. For the Research Model, the median (IQR) mortality risk prediction was 0.103 (0.048 to 0.227) and the modal prediction was 0.042.

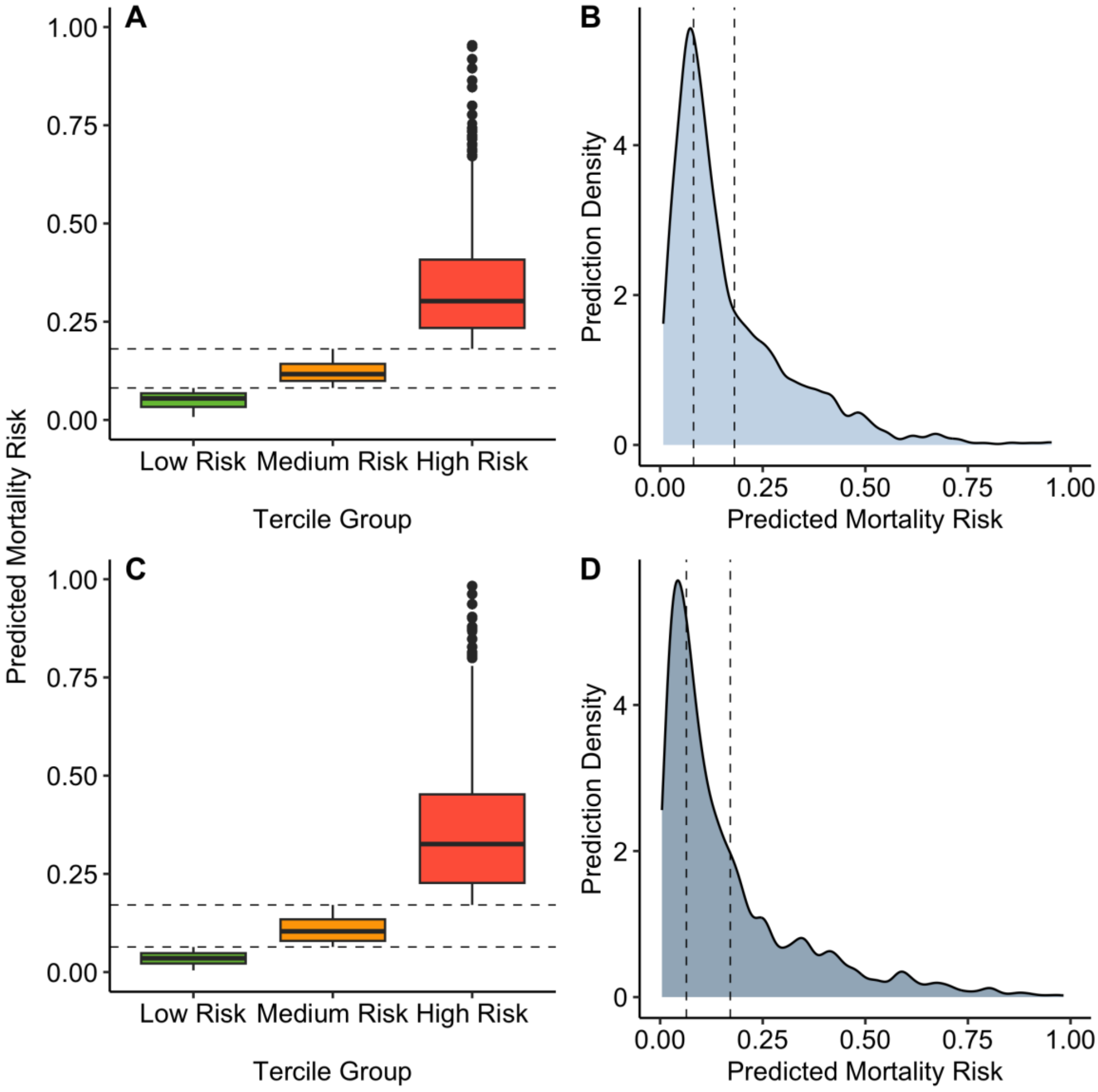

***Supplementary Table 11: Ten-week mortality by Treatment Arm and Risk Tercile***

Table showing 10-week mortality, stratified by risk tercile as defined by the Basic and Research Models. Deaths are reported for the Fluconazole + Flucytosine arm of the ACTA trial (Oral regimen), the single dose liposomal Amphotericin B arm (Ambition regimen) of the Ambition trial, and the 1-week Amphotericin B + Flucytosine arm of each of their respective trials. The Oral regimen in the ACTA trial and the Ambition regimen arm of the Ambition trial are labelled Intervention, and the 1-week Amphotericin B + Flucytosine arm is labelled standard of care (SOC). Deaths, mortality differences and hazard ratios are compared between the intervention (Oral regimen or Ambition regimen) and the standard of care for that trial and reported stratified by model and risk tercile. Deaths are described directly from the data and exclude patients for whom a risk category could not be attributed due to missing data. Mortality Difference and Hazard Ratios were calculated using multiply imputed data to account for missingness of predictor variables.

| | Deaths | | Mortality Difference | | Hazard Ratio | |
|---|---|---|---|---|---|---|
| | SOC[1,2] | Intervention[2] | Intervention v SOC[1,3] | p value | Intervention v SOC[1,3] | p value |
| **ACTA - Oral Regimen - Basic[4]** | | | | | | |
| Low Risk | 7/52 (13.5%) | 7/47 (14.9%) | 1.6% (-15.9-19) | 0.9 | 1.13 (0.41-3.14) | 0.8 |
| Medium Risk | 10/36 (27.8%) | 22/86 (25.6%) | -1.4% (-18.6-15.8) | 0.9 | 0.89 (0.43-1.85) | 0.8 |
| High Risk | 9/21 (42.9%) | 42/76 (55.3%) | 12.5% (-8.8-33.7) | 0.2 | 1.5 (0.73-3.06) | 0.3 |
| **ACTA - Oral Regimen - Research[4]** | | | | | | |
| Low Risk | 6/39 (15.4%) | 8/41 (19.5%) | 3.9% (-14-21.8) | 0.7 | 1.28 (0.45-3.63) | 0.6 |
| Medium Risk | 5/29 (17.2%) | 13/71 (18.3%) | 3.9% (-13.7-21.5) | 0.7 | 1.22 (0.48-3.11) | 0.7 |
| High Risk | 13/29 (44.8%) | 44/72 (61.1%) | 14% (-3.8-31.8) | 0.1 | 1.53 (0.85-2.76) | 0.2 |
| **Ambition – Ambition Regimen - Basic[4]** | | | | | | |
| Low Risk | 32/183 (17.5%) | 21/178 (11.8%) | -5.8% (-14.4-2.9) | 0.2 | 0.65 (0.38-1.13) | 0.1 |
| Medium Risk | 33/120 (27.5%) | 29/127 (22.8%) | -4.7% (-15.2-5.7) | 0.4 | 0.81 (0.49-1.33) | 0.4 |
| High Risk | 50/99 (50.5%) | 51/99 (51.5%) | 1% (-10.6-12.6) | 0.9 | 1.05 (0.71-1.54) | 0.8 |
| **Ambition – Ambition Regimen - Research[4]** | | | | | | |
| Low Risk | 29/181 (16%) | 17/168 (10.1%) | -6% (-14.7-2.7) | 0.2 | 0.61 (0.33-1.11) | 0.1 |
| Medium Risk | 33/122 (27%) | 30/134 (22.4%) | -4.8% (-14.9-5.3) | 0.4 | 0.82 (0.5-1.34) | 0.4 |
| High Risk | 53/99 (53.5%) | 54/102 (52.9%) | -0.5% (-11.9-10.8) | 0.9 | 1 (0.68-1.45) | 1.0 |

[1]SOC = Standard of Care

[2]Brackets indicate proportions

[3]Brackets indicate 95% confidence intervals

[4]Risk groups were defined by terciles of predicted risk in the pooled dataset. Thresholds of 8.1% and 18.1% delineated Low-Medium and Medium-High risk respectively in the basic treatment model. Thresholds of 6.4% and 17.1% delineated Low-Medium and Medium-High risk respectively in the research treatment model.
